# Methods for identifying culprit drugs in cutaneous drug eruptions: A scoping review

**DOI:** 10.1101/2021.05.11.21257038

**Authors:** Reetesh Bose, Selam Ogbalidet, Mina Boshra, Alexandra Finstad, Barbara Marzario, Christina Huang, Simone Fahim

**Affiliations:** University of Ottawa, Division of Dermatology, 737 Parkdale Avenue, Ottawa, ON K1Y 4M9, Canada; The Ottawa Hospital Division of Dermatology, 737 Parkdale Avenue, Ottawa, ON K1Y 4M9, Canada; University of Ottawa Faculty of Medicine, 451 Smyth road, Ottawa, ON, K1H8L1, Canada

**Keywords:** Drug rash, cutaneous adverse reaction, culprit drug identification, algorithm, causality assessment, hypersensitivity

## Abstract

**Background:** Cutaneous drug eruptions are a significant source of morbidity, mortality, and cost to the healthcare system. Identifying the culprit drug is essential; however, despite numerous methods being published, there are no consensus guidelines.

**Objectives:** Conduct a scoping review to identify all published methods of culprit drug identification for cutaneous drug eruptions, compare the methods, and generate hypotheses for future causality assessment studies.

**Eligibility criteria:** Peer-reviewed publications involving culprit drug identification methods.

**Sources of evidence:** Medline, Embase, and Cochrane Central Register of Controlled Trials.

**Charting methods:** Registered PRISMA-ScR format protocol on Open Science Forum.

**Results:** In total, 135 publications were included comprising 656,635 adverse drug events, most of which were cutaneous. There were 54 methods of culprit drug identification published, categorized as algorithms, probabilistic approaches, and expert judgment.

Algorithms had higher sensitivity and positive predictive value, but lower specificity and negative predictive value. Probabilistic approaches had lower sensitivity and positive predictive value, but higher specificity and negative predictive value. Expert judgment was subjective, less reproducible, but the most frequently used to validate other methods. Studies suggest that greater accuracy may be achieved by specifically assessing cutaneous drug eruptions and using combinations of causality assessment categories.

**Conclusions:** Culprit drug identification for adverse drug reactions remains a challenge. Many methods have been published, but there are no consensus guidelines. Using causality assessment methods specifically for cutaneous drug eruptions and combining aspects of the different causality assessment categories may improve efficacy. Further studies are needed to validate this hypothesis.

## INTRODUCTION

### Rationale

Skin reactions are the most common manifestation of adverse drug events (ADEs). Cutaneous drug eruptions (CDEs) are a common and a significant source of morbidity and mortality. (1) The mainstay of management is identifying and stopping the culprit drug. (2) Many methods have been proposed for culprit drug identification (CDI), but presently there are no consensus guidelines. (2–4) There are few causality assessment methods (CAMs) that are specific to CDEs, but most methods are applied to all types of ADEs.

There are 3 main categories of causality assessment: operational algorithms, probabilistic approaches, and expert judgment. Each of the published methods of culprit drug identification usually fit into one of these categories. (4,5) Operational algorithms involve specific criteria related to drug exposure and the resulting adverse event. They are relatively simple to use and are reproducible, but can be restrictive because specific information is often required. (6) Probability assessment methods use likelihood ratios relating to the case. For example, different types of CDEs often have specific latencies between drug exposure and rash onset, which narrows the time-frame when the drug exposure likely occurred. Amongst other criteria, the frequency with which the drug has been reported to cause the adverse event is expressed as an overall probability score, implicating the drug with the highest score as the culprit drug. (7,8) Unfortunately, such methods often rely on previously reported adverse events. Probabilistic methods can be complex, time consuming, do not account for unreported drugs, and are not always practical in real-world settings. (6,9) Expert judgment approaches are based on the clinical opinion of an expert physician, the physician treating the patient, or sometimes a clinical pharmacist. All available data in a case is considered, and how much weight each piece of data holds is determined by the expert and a decision regarding causality is made based on this judgment. Although this is one of the simplest methods, it is often subjective and biased, with poor inter-rater agreement and reproducibility. (6,10)

ADEs are heterogeneous, and often affect more than one organ system. Clinical signs of drug eruptions have been extensively studied and categorized. Cutaneous manifestations often happen early and may allow for a more accurate timeline compared to internal involvement which can often be sub-clinical. Developing CAMs using ADE databases may not always be accurate because of the heterogeneity between drug exposures, cutaneous and systemic manifestations. A scoping review was employed to identify all published methods of CDI for CDEs, compare the methods to uncover strengths and limitations, and generate hypotheses for future causality assessment studies.

## METHODS

### Protocol and Registration

The protocol was drafted using the Preferred Reporting Items for Systematic Reviews and Meta-analysis Protocols extension for scoping reviews (PRISMA-ScR) (11), and was registered with Open Science Forum (OSF). (Bose R, Boshra M, Ogbalidet S, Finstad A, Fahim S. A scoping review of cutaneous drug eruptions and methods for identifying culprit drugs. 2020. osf.io/r7k3z.)

### Eligibility criteria

Inclusion Criteria: Peer-reviewed publications were included if they were written in English, published in 1993 or later, and involved a method of CDI. A cut-off of 1993 was selected because notable CDE reclassifications had taken place prior to this time, specifically the classification of Stevens Johnson syndrome as a separate entity from erythema multiforme. Relevant review articles were analyzed separately from articles containing primary patient data.

Exclusion Criteria: Publications were excluded if there was no mention of a method for identifying culprit drugs, if all participants were ≤18 years old, or if the publication was of low-level evidence (i.e. case reports, case series involving <10 cases, commentaries, editorials, conference posters/abstracts or opinion pieces).

### Information sources

Medline (Ovid), Embase, and Cochrane Central Register of Controlled Trials (CENTRAL) databases were used. The most recent search was executed: February 5^th^, 2020. Studies were imported into Covidence, a systematic review software.

### Search strategy

Developed and conducted with the assistance of our institutional research library services. See Table 1 for details.

**Table 1.**
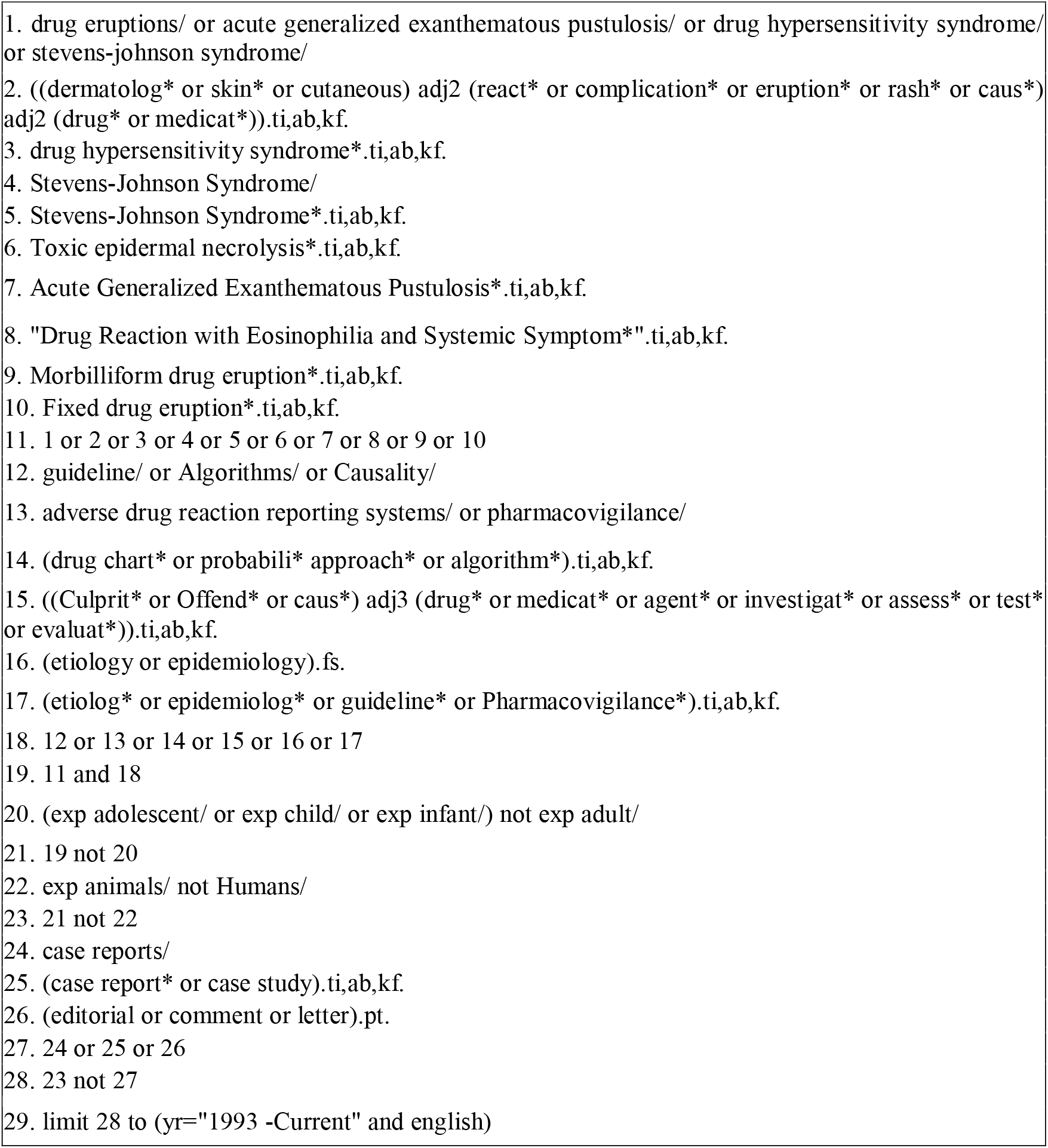
The following electronic search strategy, developed with assistance of a research librarian, was conducted with Medline (Ovid), Embase, and Cochrane Central Register of Controlled Trials (CENTRAL) databases.

### Selection of sources of evidence

A list of publications was generated with the search strategy, and duplicates were removed. Two groups of reviewers independently screened all abstracts followed by full texts. Disagreements were settled by consensus vote. Reference lists of included publications were screened for relevant studies not captured by the initial search. Publications involving patients were separated from review articles (See Figure 1).

**Figure 1.**
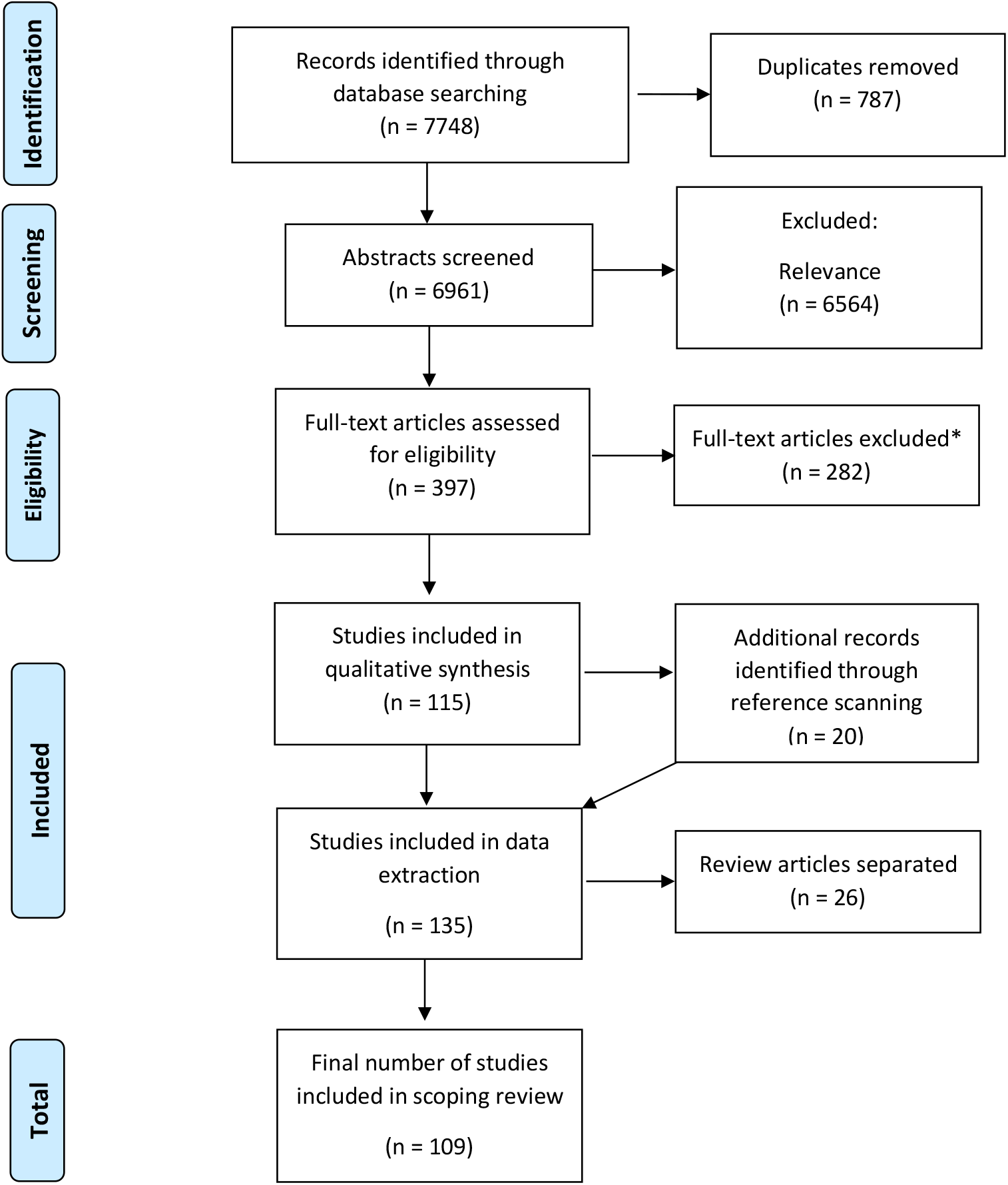
PRISMA-ScR flow diagram mapping the process used to include and exclude publications. *Excluded studies (n=282): Wrong study design (n=106), wrong outcomes (n=89), abstract only (n=28), reviews with causality testing not included (n=16), wrong patient population (n=12), wrong setting (n=9), pediatric only (n=7), posters (n=6), duplicates (n=5), unavailable (n=2) and not available in English (n=2).

### Data charting process

*A priori* data categories were used to organize extracted data. Review articles were analyzed separately to determine if additional data categories should be extracted from the primary studies containing patient data. Quality assurance assessment for accuracy of entered data was conducted by the primary author.

### Data items extracted

The list of data extraction items can be found in Table 2. The data from included publications were organized into tabular format using Microsoft Excel.

**Table 2.** A priori data extraction items for included publications in the scoping review.

|  |
| --- |
| • Year published, author, country of publication, patient age, and gender predominance<br>• Number of cases included in study<br>• Cutaneous adverse reaction type(s) recorded<br>• Culprit drug(s) identified for each drug eruption type<br>• Latency between drug exposure and onset of rash<br>• Risk factors and associated conditions<br>• Method used to rule out other causes of rash<br>• Drug cross-reactions, if indicated<br>• Viral reactivation if indicated (e.g. HHV6& 7 for DRESS)<br>• Tissue biopsy if performed to confirm eruption type<br>• Causality assessment method used<br>• In vitro investigations for culprit drug identification, if performed<br>• Confirmatory/provocation testing, if performed<br>• Quantitative/qualitative analysis comparing drug eruptions types<br>• Statistical methods to analyze methods of culprit drug identification<br>• Type of study and level of evidence |

### Synthesis of results

The extracted data from each publication was entered into a database for easier synthesis and comparison of data (Supplemental Table 4). Some CAMs used different syntax to assign a likelihood rating for suspected culprit drugs. Analogous terms were grouped together to compare the different CAMs (Table 5). Most studies used a Likert-style scale to assign different levels of causality for suspected culprit drugs being investigated. For example, for 5-term scales, causality assessment *Method A* may report the likelihood of a culprit drug as: very likely, likely, possible, unlikely, or very unlikely. *Method B* may report likelihood as: certain, probable, possible, doubtful, or exclude. In this case the analogous categories were grouped: ‘Certain/very likely’, ‘probable/likely’, ‘possible’, ‘doubtful/unlikely’, and ‘exclude/very unlikely’. Some methods used a 3 term rating scale such as ‘positive’, ‘neutral’, and ‘negative’. To report this 3-term scale with the 5-term scales the ‘positive’ rating would be grouped with ‘very likely/certain’, the ‘neutral’ rating would be grouped with ‘possible’ and the ‘negative’ rating would be grouped with ‘exclude/very unlikely’.

**Table 3:** Characteristics of included papers (N=135; 109 studies, 26 reviews). ADE: adverse drug events, SJS/TEN: Stevens Johnson syndrome/toxic epidermal necrolysis, MDE: morbilliform drug eruption, FDE: fixed drug eruption, Drug rash eosinophilia and systemic symptoms.

| Parameter | Measure |
| --- | --- |
| Publication types included | 135 (109 studies; 26 reviews) |
| Year of publication range | 1993-2020 |
| Country of publication | 15 different countries:<br>• India 34 (31.2%)<br>• Europe 24 (22.0%)<br>• Israel 8 (7.3%)<br>• North America 6 (5.5%)<br>• South America 6 (5.5%)<br>• China 5 (4.6%)<br>• Singapore/Philippines/Malasia 4 (3.7%)<br>• Other: 22 (20.2%) |
| Age range of patients | 0-95 |
| Gender predominance | Female 58 (53.2%) |
| Total number of ADE cases | 656,635 |
| Study types: | • Retrospective 81 (74.3%)<br>• Prospective 27 (24.8)<br>• Both 1 (0.9%) |
| Types of cutaneous eruption<br>• Number of studies (%) | 33 different types:<br>• SJS/TEN 69 (63.3%)<br>• MDE 39 (35.8%)<br>• FDE 31 (29.1%)<br>• DRESS 32 (29.1%)<br>• Urticaria 24 (22.0%) |
| Tissue biopsy performed | 27 (24.8%) |
| Causality assessment methods | 54 different methods |
| Causality assessment categories | • Operational algorithmic 34 (63.0%)<br>• Probabilistic 18 (33.3%)<br>• Expert judgment: 2 (3.7%) |
| Lab-based methods | • 9 methods<br>• 26 publications: 11 studies, 14 reviews |
| Studies comparing methods | 29 (26.6%) |
| Studies with statistical analyses | 19 (17.3%) |
| Confirmatory testing performed | 21 (19.3%) |

**Table 4.** Raw extracted data form. Supplemental. Excluded from pre-print.

**Table 5.**
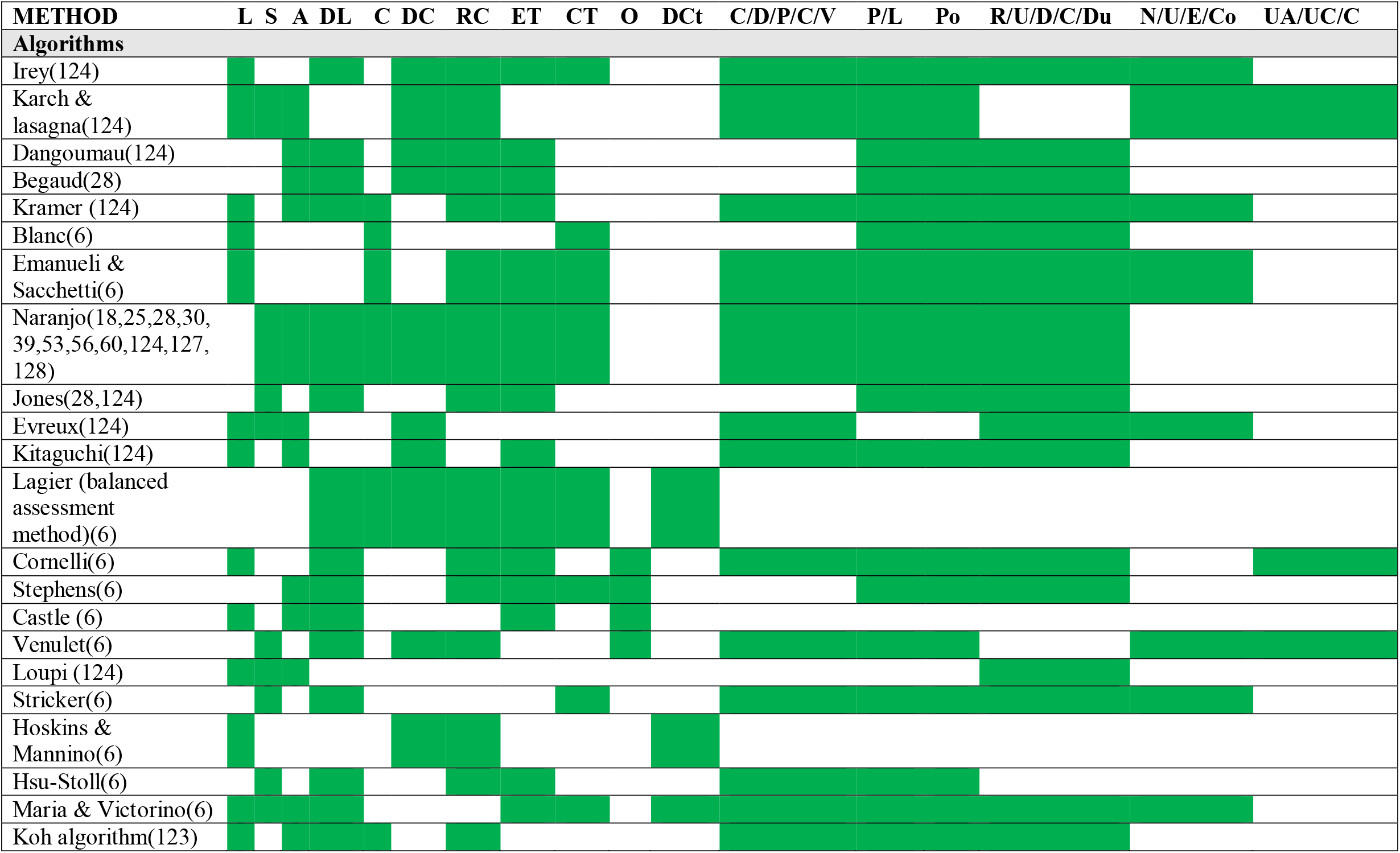
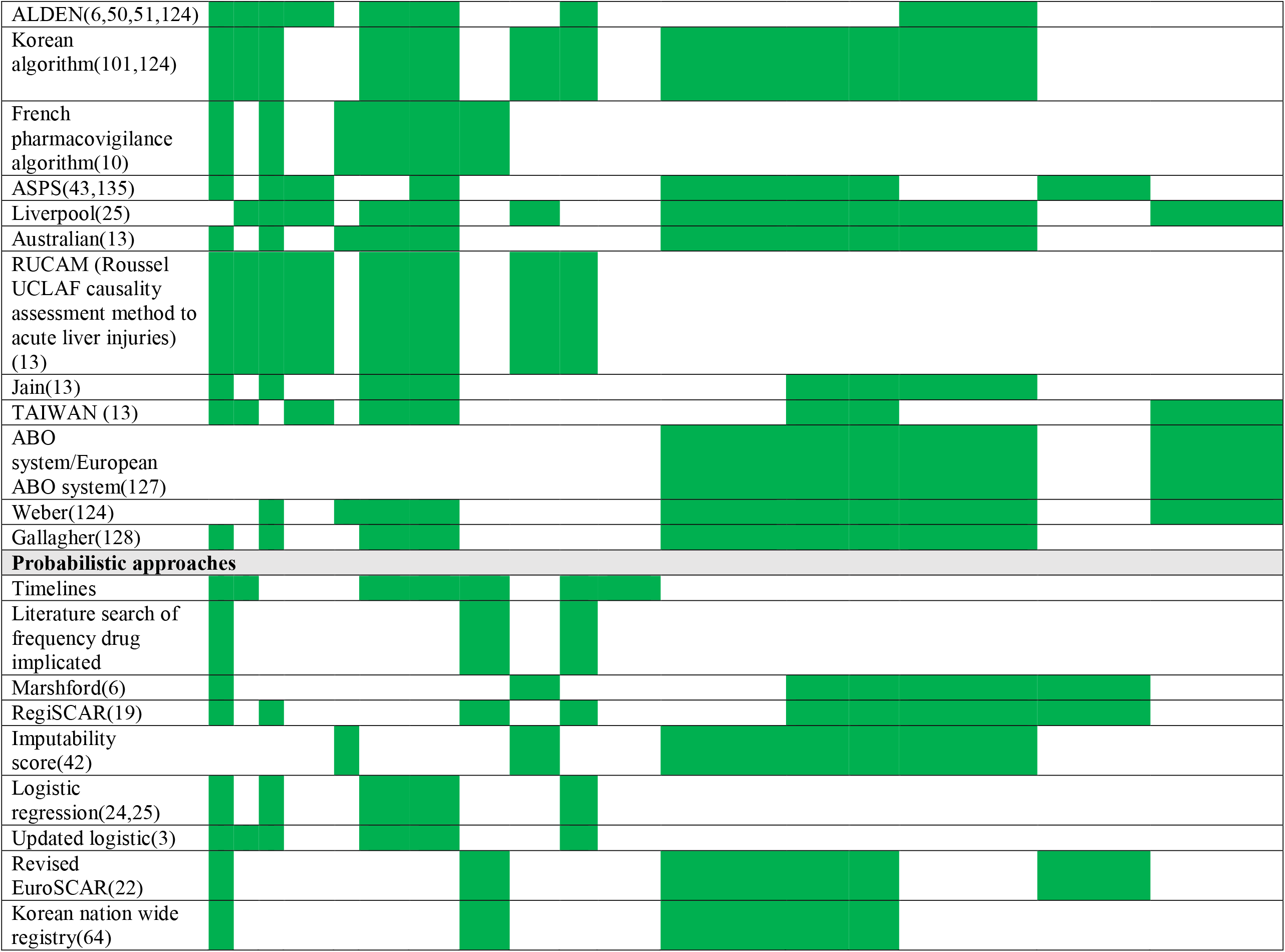
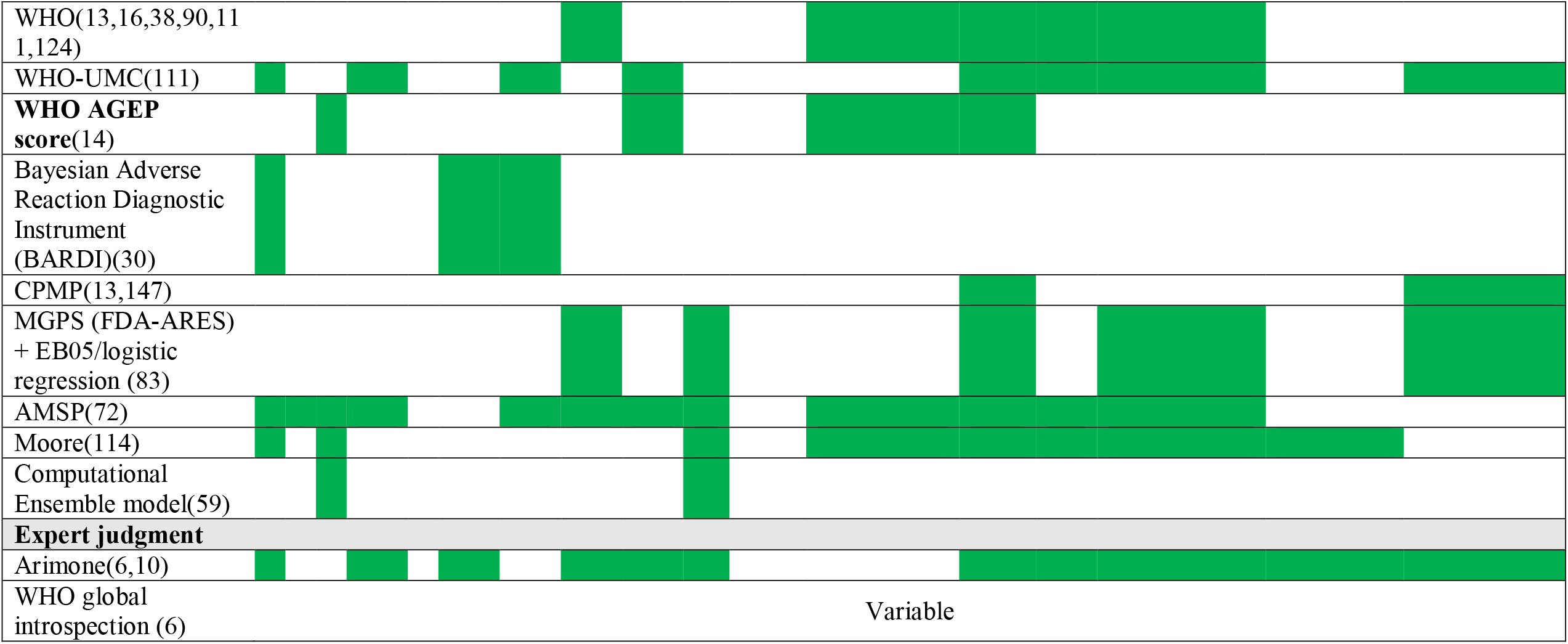
Analysis and comparison of criteria/components of published methods of culprit drug identification. Green = criteria/component present, White = criteria/component not present in method. L: latency; S: prior sensitization; A: alternate diagnosis; DL: drug level/dose change; C: challenge, DC: de-challenge, RC: re-challenge; ET: eruption type; CT: confirmatory testing; O : other drug suspect; DCt: Drug chart; ASPS: Spanish pharmacovigilance algorithm system C/D/P/C/V: certain, definite, positive, causative, very likely; P/L: probable/likely; Po: possible; R/U/D/C/Du: remote/unlikely/doubtful/coincidental/dubious; N/U/E/Co: negative, unrelated, exclude, contradictory; Ua/Uc/C: unaccessable/unclassifiable/conditional.

For the studies reporting quantitative results such as sensitivity and specificity (See table 6), the data analyzed for this study was reported in a descriptive manner in line with accepted reporting guidelines for scoping reviews.(11)

**Table 6.**
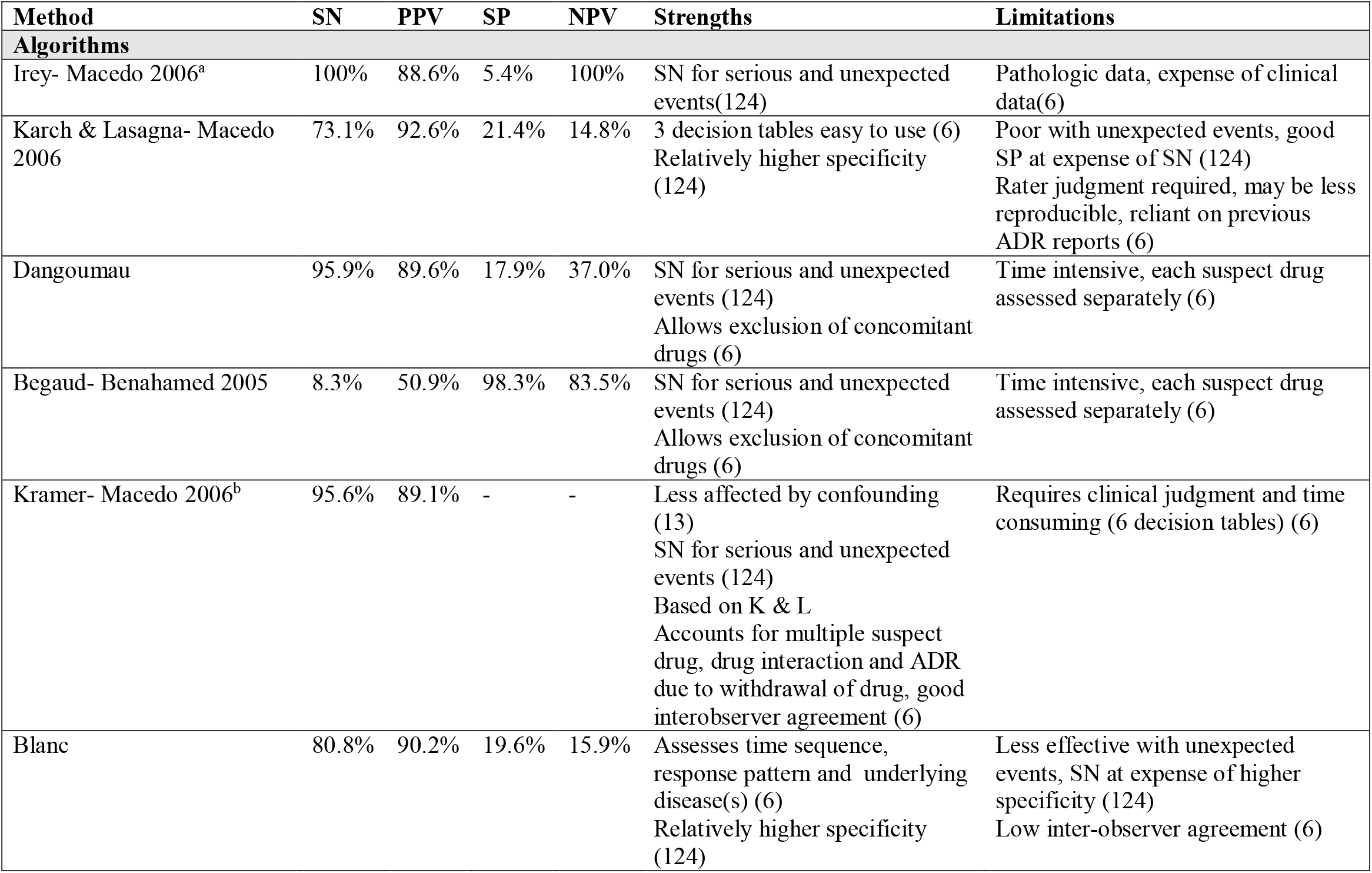
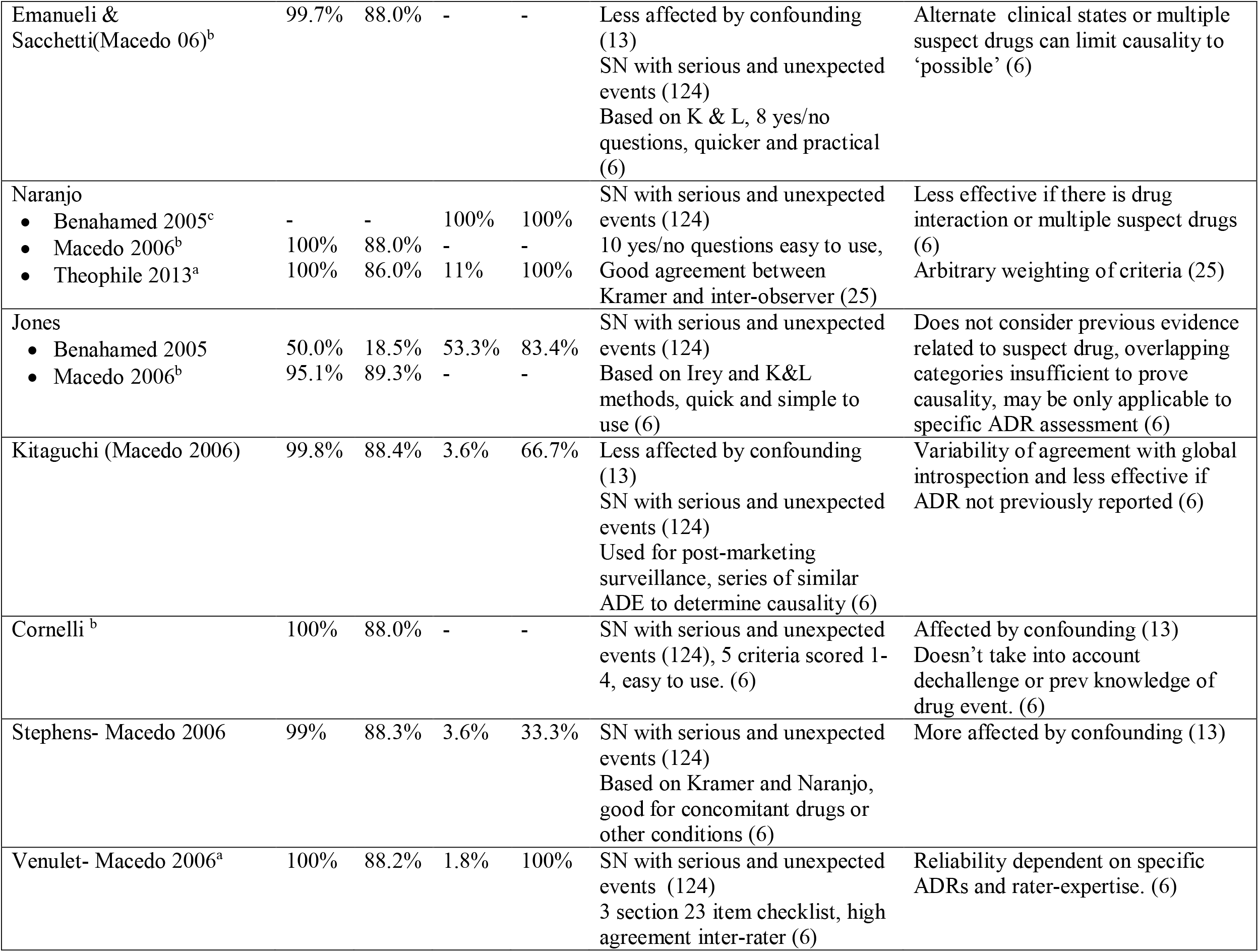
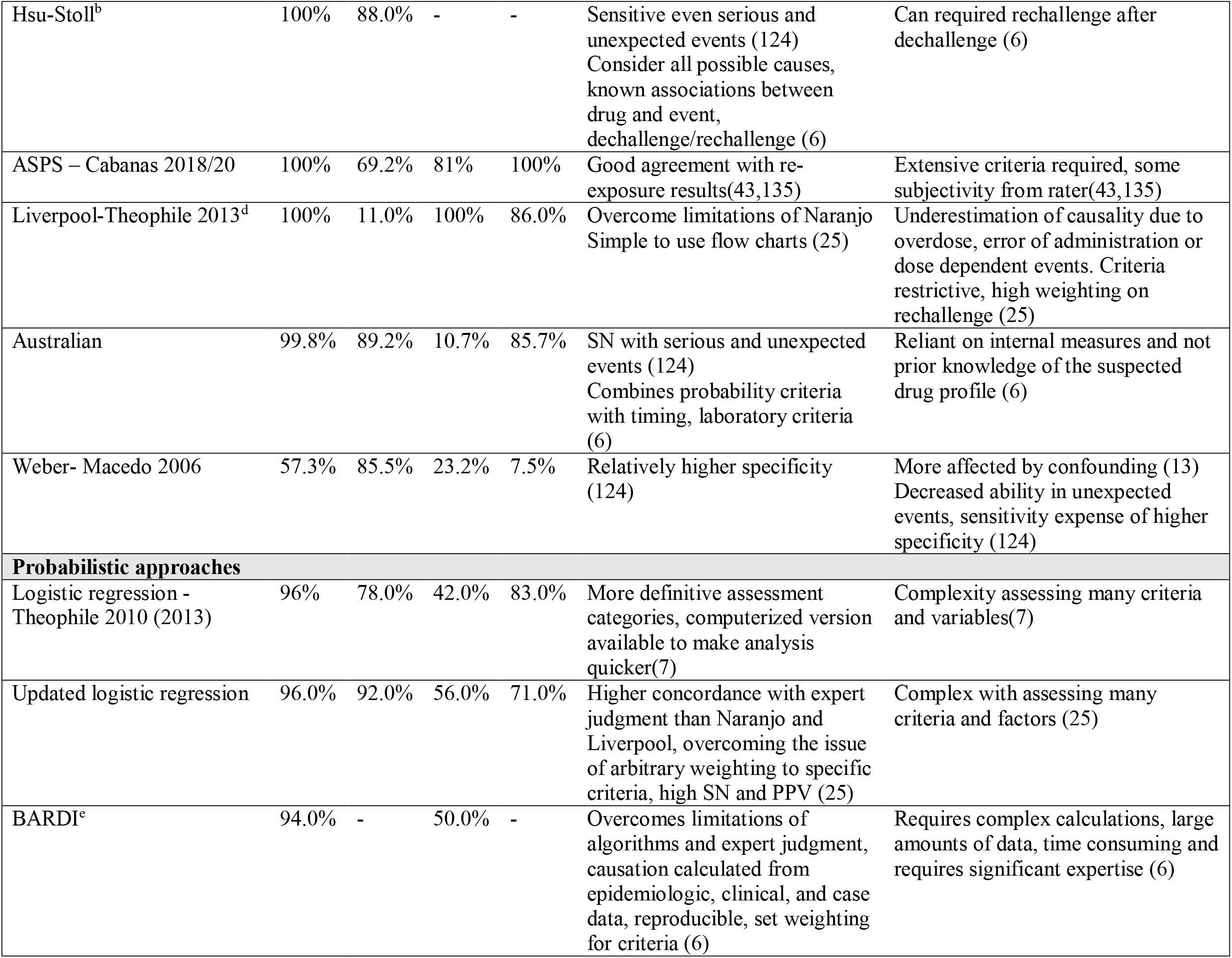
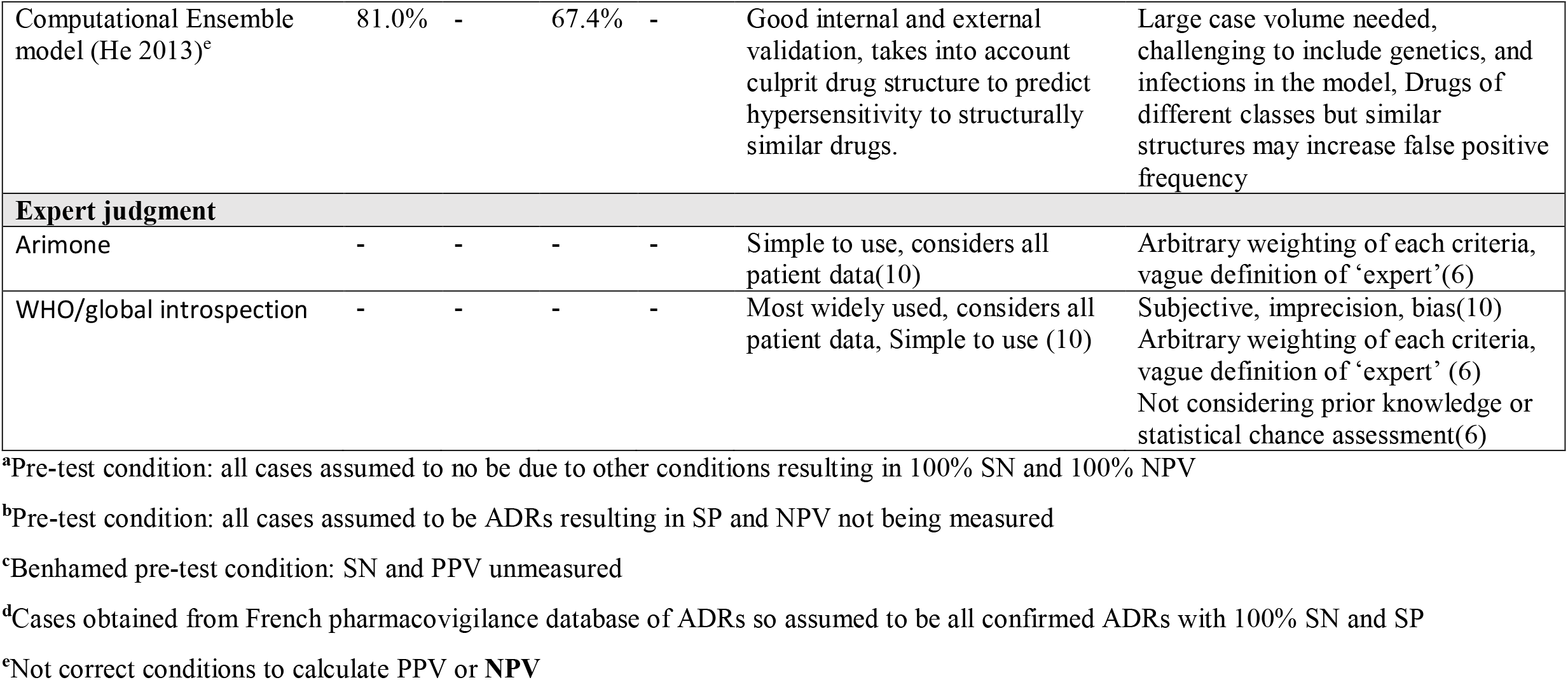
Published causality assessment methods that have been quantitatively studied. SN: sensitivity, PPV: positive predictive value, SP: specificity, NPV: negative predictive value, *SPVA*: Spanish pharmacovigilance algorithm, BARDI: Bayesian adverse reaction diagnostic instrument. Cornelli, Emanueli, Hsu Stoll, Jones, Kramer and Naranjo algorithms assumed all the identified adverse events were ADRs therefore presenting 0% specificity and 0% NPV. Given (wider 95% CIs) as a result of the small number of algorithm’s high sensitivity, even for serious or uncases considered non-drug related by the GI.

Review articles were analyzed to ensure all necessary data items were used to extract data from primary studies and to compare the findings and associations uncovered in this review.

## RESULTS

### Selection of sources of evidence

Sources of evidence were gathered using the search strategy, registered on OSF (Table 1). The screening process, and application of inclusion and exclusion criteria was conducted as per the PRISMA-ScR extension protocol and reasons for exclusion of sources of evidence were recorded (Figure 1).

### Characteristics of sources of evidence

The characteristics of included sources of evidence were recorded using the items outlined in Table 2 and shown in Table 3. Studies were primarily retrospective cohort, case-control, and prospective cohort or observational.

#### Results of individual sources of evidence

For the complete list of publications and extracted data included in the scoping review (Table 4, supplemental).

### Synthesis of results

A total of 109 peer-reviewed articles with patient data involving at least one CDI method, were analyzed (raw data, Table 4, study characteristics Table 3). There were 26 review articles that were analyzed to compare parameters and determine the relevance in the present review (Table 7). Overall, data from 656,635 case events were represented. A total of 29 papers (26.6%) compared 2 or more CAMs, and 19 (17.4%) statistically analysed the methods. There were 54 different CAMs published and operational algorithms were the most studied (71 publications, 65.1%). From these 71 publications, 34 CAMs were identified as operational algorithms. There were 58 publications (53.2%) that studied probabilistic approaches in which 18 CAMs were identified. Finally, 48 publications (44.0%) involved the use of expert judgment and 2 different methods were identified. Many studies used more than one CAM.

**Table 7.**
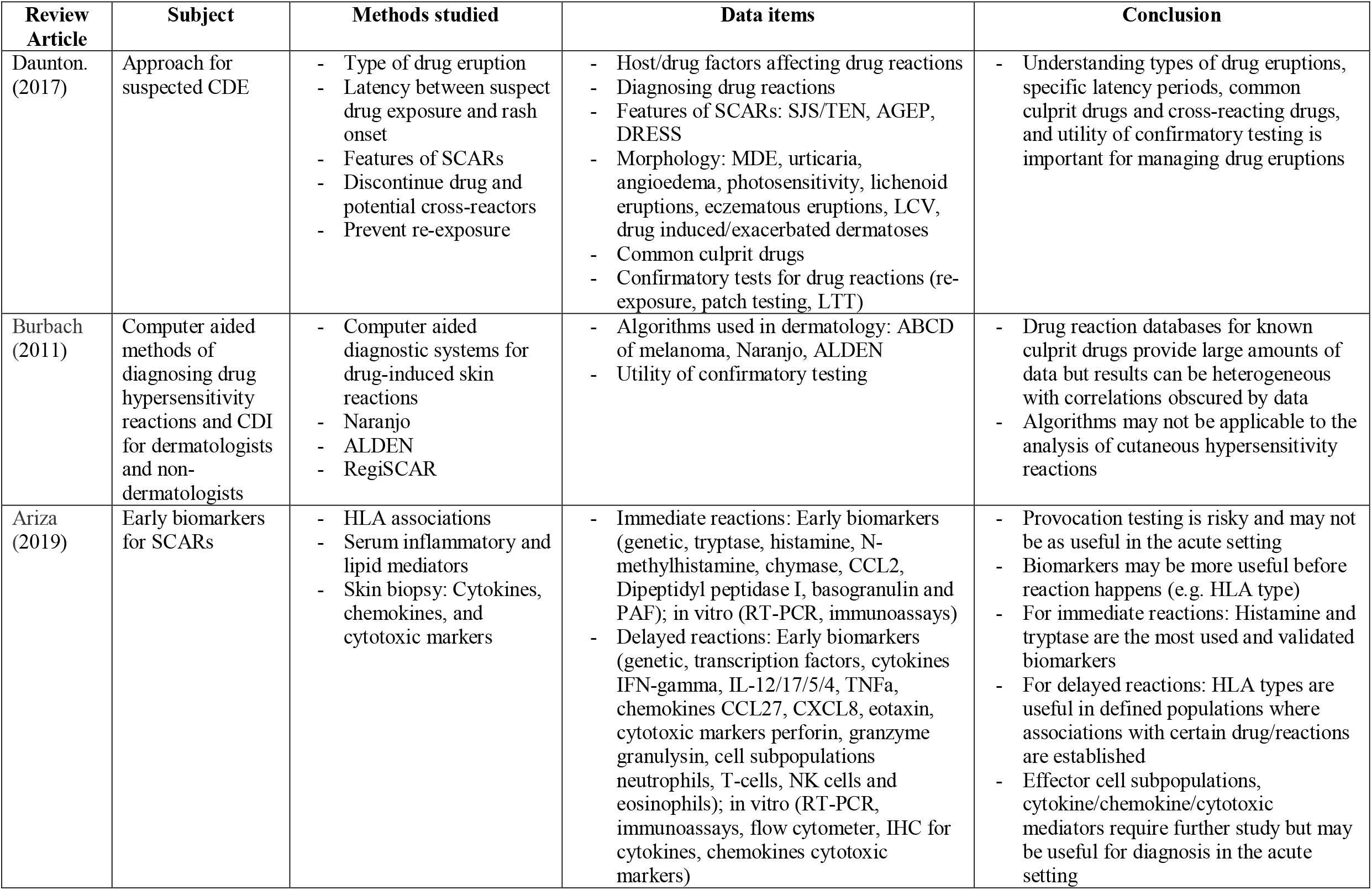
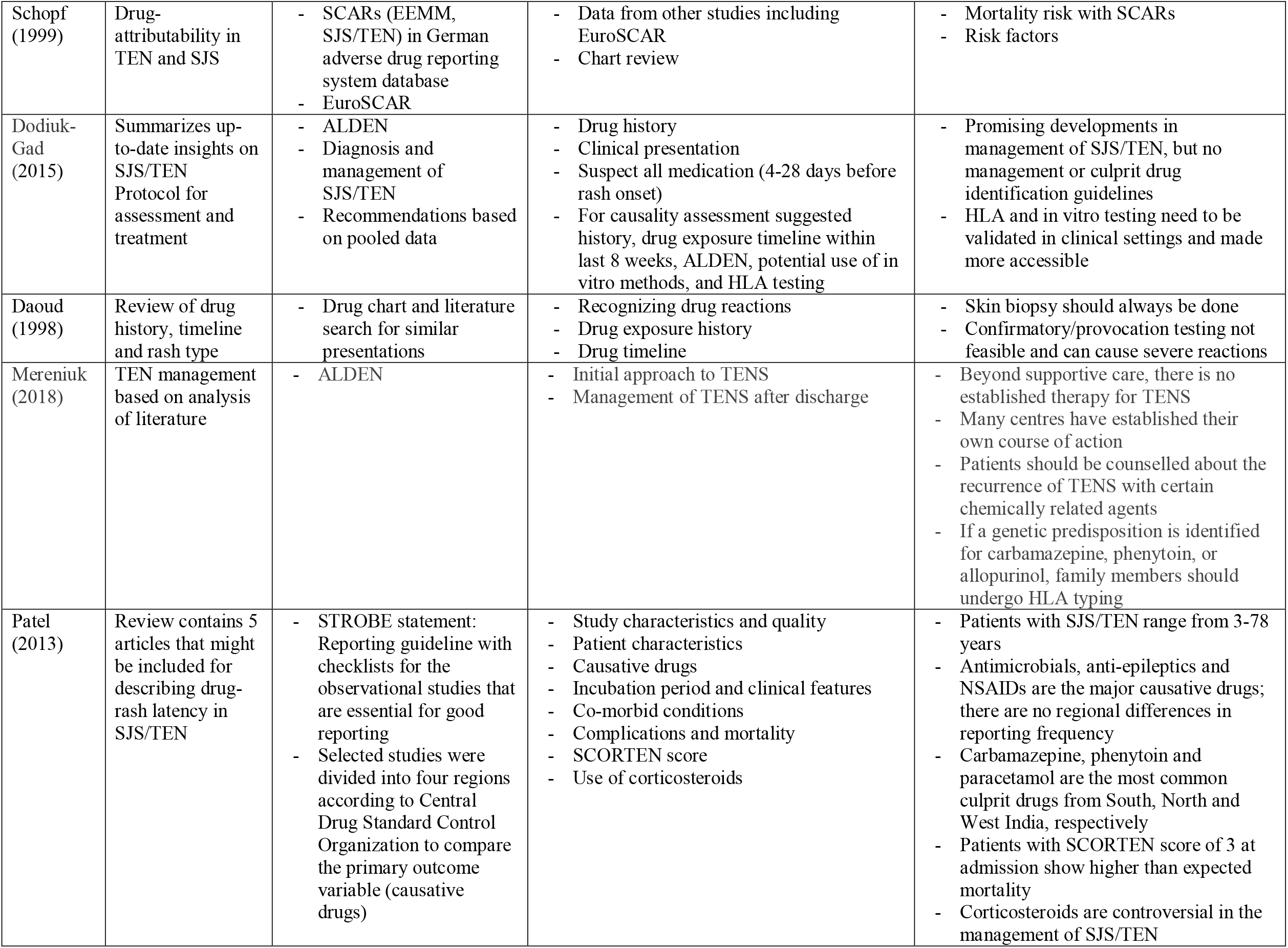
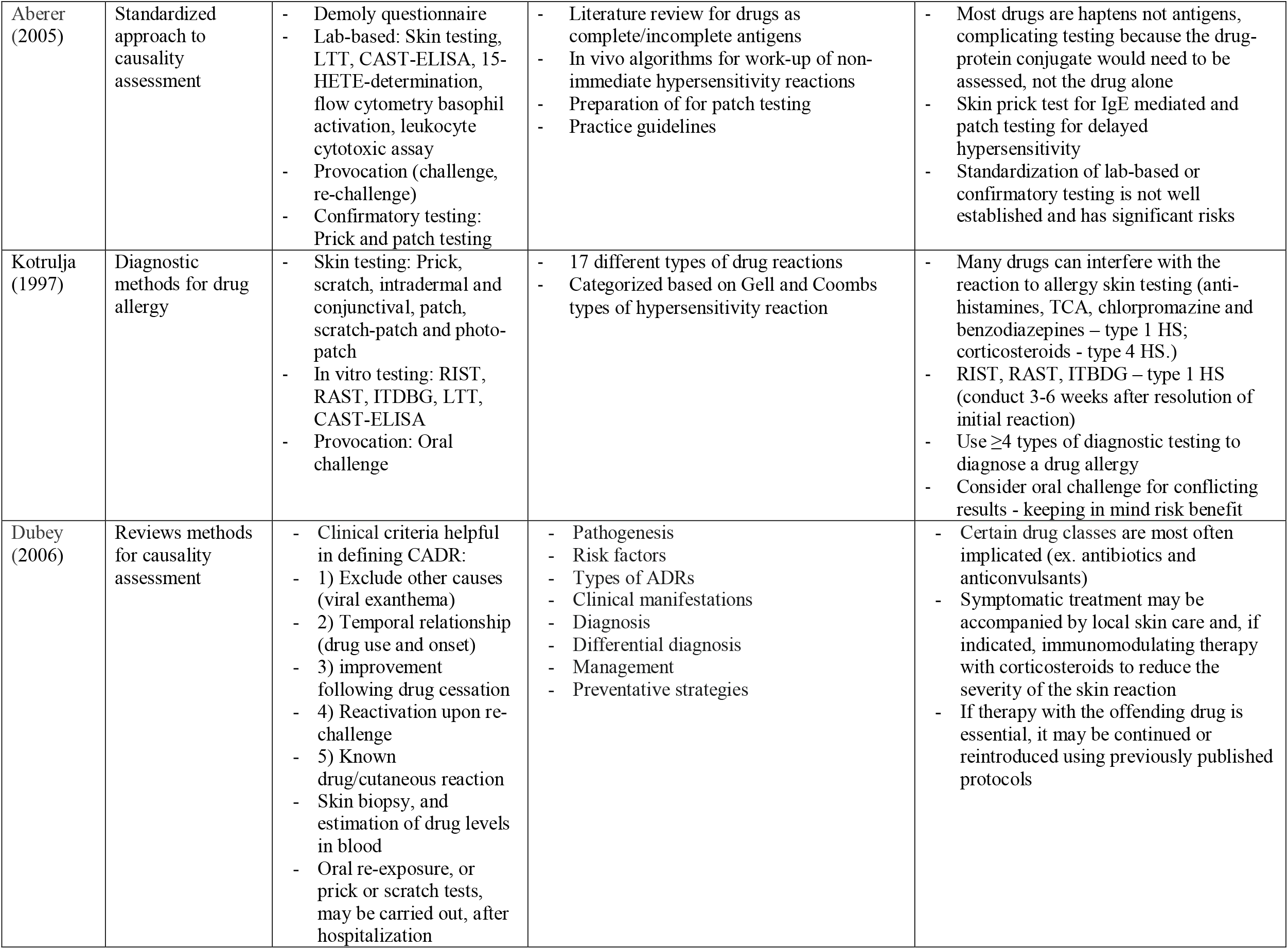
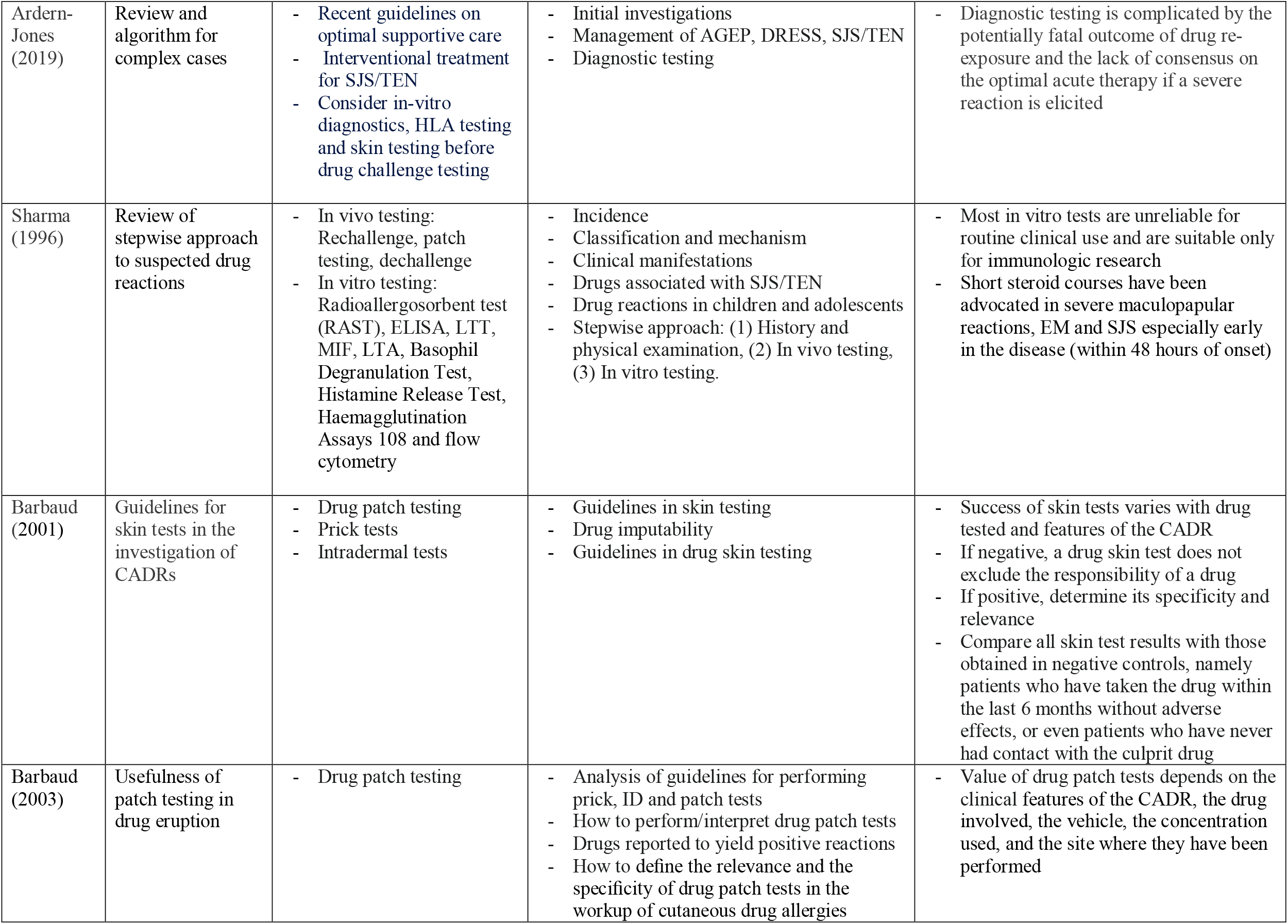
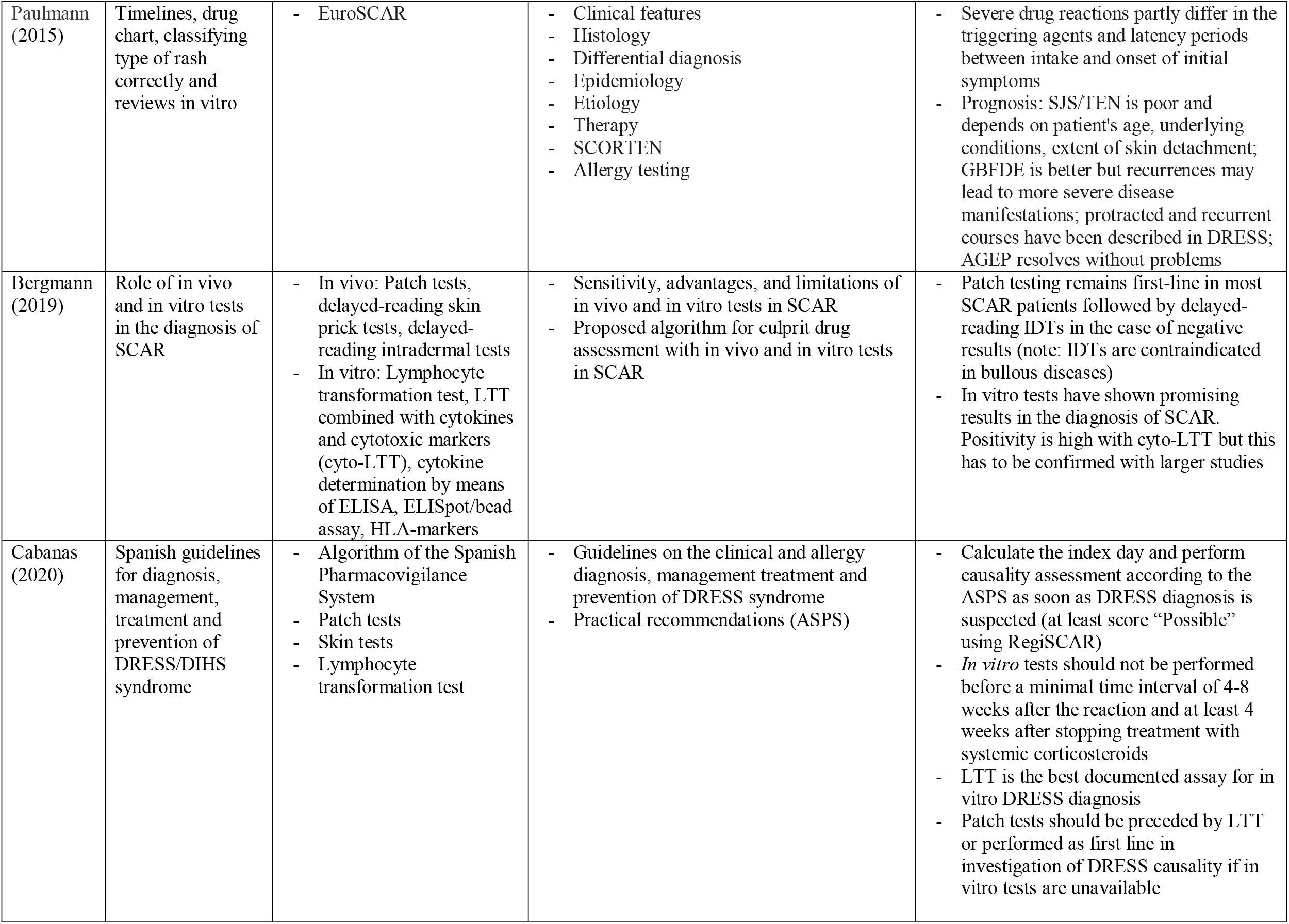
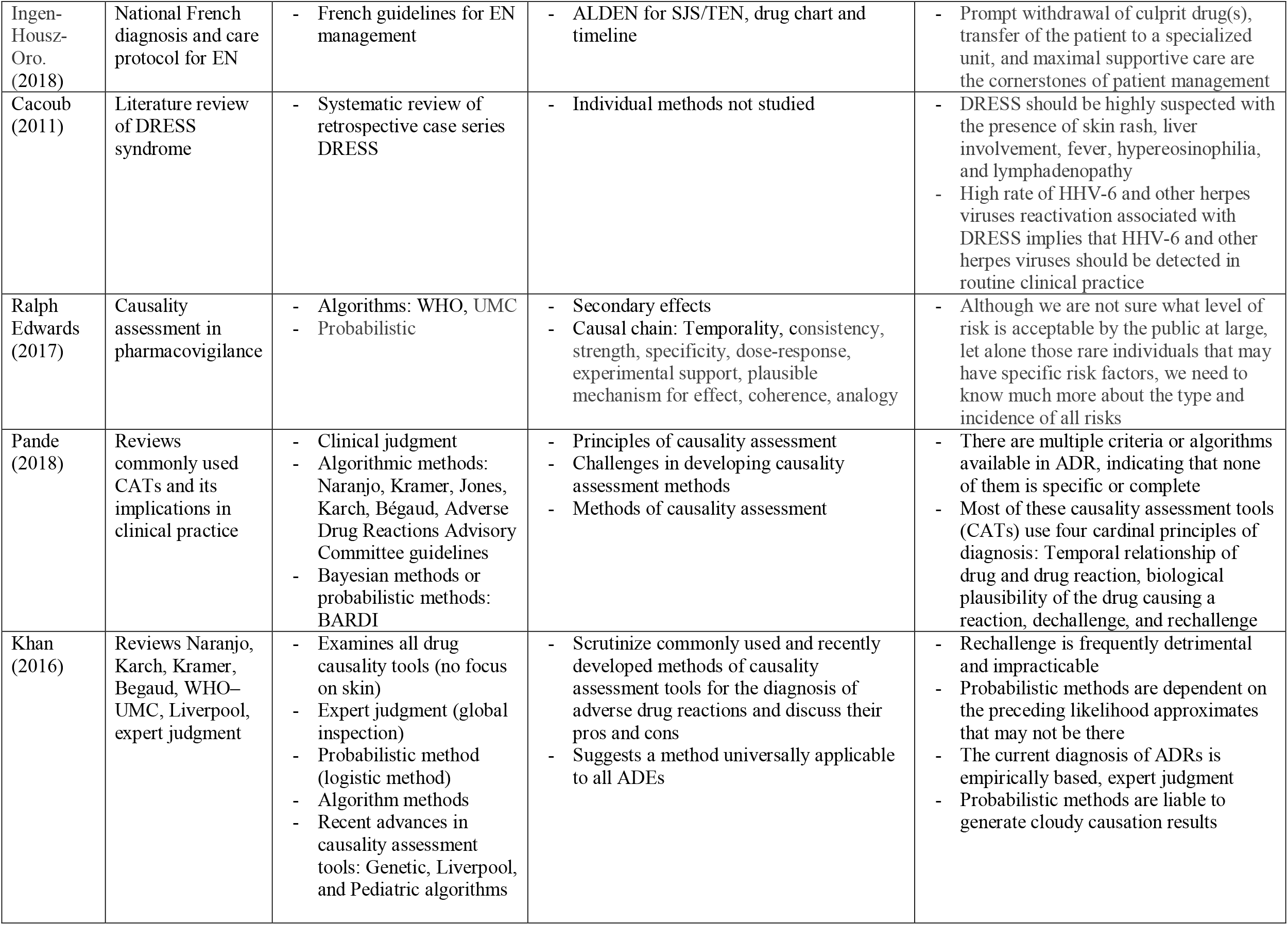
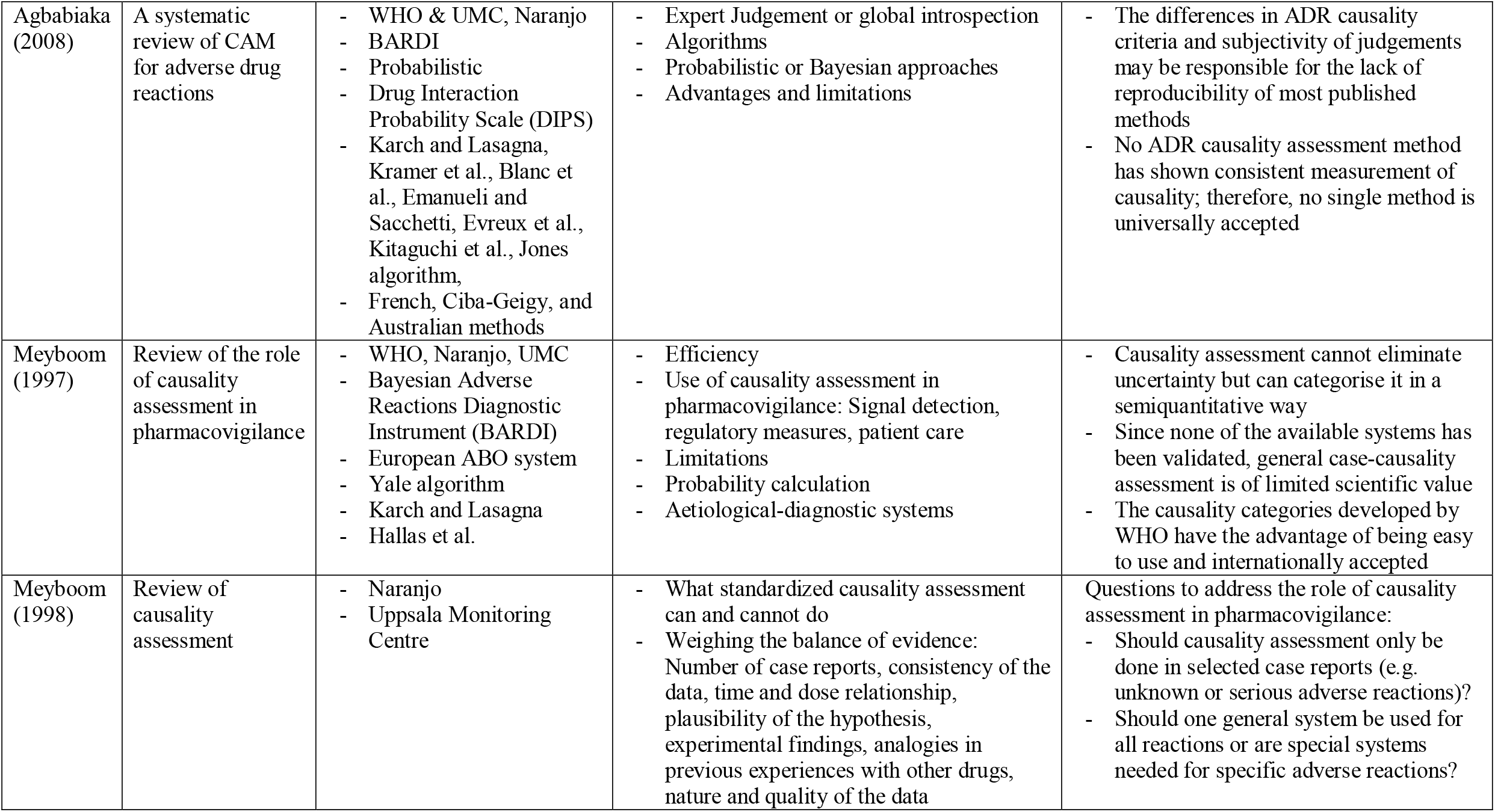
Analysis of review article publications included in study. CDE: cutaneous drug eruptions; CDI: culprit drug identification; CAM: causality assessment methods; EEM: erythema exsudativum multiforme majus.

| Review Article | Subject | Methods studied | Data items | Conclusion |
| --- | --- | --- | --- | --- |
| Daunton. (2017) | Approach for suspected CDE | - Type of drug eruption<br>- Latency between suspect drug exposure and rash onset<br>- Features of SCARs<br>- Discontinue drug and potential cross-reactors<br>- Prevent re-exposure | - Host/drug factors affecting drug reactions<br>- Diagnosing drug reactions<br>- Features of SCARs: SJS/TEN, AGEP, DRESS<br>- Morphology: MDE, urticaria, angioedema, photosensitivity, lichenoid eruptions, eczematous eruptions, LCV, drug induced/exacerbated dermatoses<br>- Common culprit drugs<br>- Confirmatory tests for drug reactions (re-exposure, patch testing, LTT) | - Understanding types of drug eruptions, specific latency periods, common culprit drugs and cross-reacting drugs, and utility of confirmatory testing is important for managing drug eruptions |
| Burbach (2011) | Computer aided methods of diagnosing drug hypersensitivity reactions and CDI for dermatologists and non-dermatologists | - Computer aided diagnostic systems for drug-induced skin reactions<br>- Naranjo<br>- ALDEN<br>- RegiSCAR | - Algorithms used in dermatology: ABCD of melanoma, Naranjo, ALDEN<br>- Utility of confirmatory testing | - Drug reaction databases for known culprit drugs provide large amounts of data but results can be heterogeneous with correlations obscured by data<br>- Algorithms may not be applicable to the analysis of cutaneous hypersensitivity reactions |
| Ariza (2019) | Early biomarkers for SCARs | - HLA associations<br>- Serum inflammatory and lipid mediators<br>- Skin biopsy: Cytokines, chemokines, and cytotoxic markers | - Immediate reactions: Early biomarkers (genetic, tryptase, histamine, N-methylhistamine, chymase, CCL2, Dipeptidyl peptidase I, basogranulin and PAF); in vitro (RT-PCR, immunoassays)<br>- Delayed reactions: Early biomarkers (genetic, transcription factors, cytokines IFN-gamma, IL-12/17/5/4, TNFa, chemokines CCL27, CXCL8, eotaxin, cytotoxic markers perforin, granzyme granulysin, cell subpopulations neutrophils, T-cells, NK cells and eosinophils); in vitro (RT-PCR, immunoassays, flow cytometer, IHC for cytokines, chemokines cytotoxic markers) | - Provocation testing is risky and may not be as useful in the acute setting<br>- Biomarkers may be more useful before reaction happens (e.g. HLA type)<br>- For immediate reactions: Histamine and tryptase are the most used and validated biomarkers<br>- For delayed reactions: HLA types are useful in defined populations where associations with certain drug/reactions are established<br>- Effector cell subpopulations, cytokine/chemokine/cytotoxic mediators require further study but may be useful for diagnosis in the acute setting |
| Schopf (1999) | Drug-attributability in TEN and SJS | - SCARs (EEMM, SJS/TEN) in German adverse drug reporting system database<br>- EuroSCAR | - Data from other studies including EuroSCAR<br>- Chart review | - Mortality risk with SCARs<br>- Risk factors |
| Dodiuk-Gad (2015) | Summarizes up-to-date insights on SJS/TEN Protocol for assessment and treatment | - ALDEN<br>- Diagnosis and management of SJS/TEN<br>- Recommendations based on pooled data | - Drug history<br>- Clinical presentation<br>- Suspect all medication (4-28 days before rash onset)<br>- For causality assessment suggested history, drug exposure timeline within last 8 weeks, ALDEN, potential use of in vitro methods, and HLA testing | - Promising developments in management of SJS/TEN, but no management or culprit drug identification guidelines<br>- HLA and in vitro testing need to be validated in clinical settings and made more accessible |
| Daoud (1998) | Review of drug history, timeline and rash type | - Drug chart and literature search for similar presentations | - Recognizing drug reactions<br>- Drug exposure history<br>- Drug timeline | - Skin biopsy should always be done<br>- Confirmatory/provocation testing not feasible and can cause severe reactions |
| Mereniuk (2018) | TEN management based on analysis of literature | - ALDEN | - Initial approach to TENS<br>- Management of TENS after discharge | - Beyond supportive care, there is no established therapy for TENS<br>- Many centres have established their own course of action<br>- Patients should be counselled about the recurrence of TENS with certain chemically related agents<br>- If a genetic predisposition is identified for carbamazepine, phenytoin, or allopurinol, family members should undergo HLA typing |
| Patel (2013) | Review contains 5 articles that might be included for describing drug-rash latency in SJS/TEN | - STROBE statement: Reporting guideline with checklists for the observational studies that are essential for good reporting<br>- Selected studies were divided into four regions according to Central Drug Standard Control Organization to compare the primary outcome variable (causative drugs) | - Study characteristics and quality<br>- Patient characteristics<br>- Causative drugs<br>- Incubation period and clinical features<br>- Co-morbid conditions<br>- Complications and mortality<br>- SCORTEN score<br>- Use of corticosteroids | - Patients with SJS/TEN range from 3-78 years<br>- Antimicrobials, anti-epileptics and NSAIDs are the major causative drugs; there are no regional differences in reporting frequency<br>- Carbamazepine, phenytoin and paracetamol are the most common culprit drugs from South, North and West India, respectively<br>- Patients with SCORTEN score of 3 at admission show higher than expected mortality<br>- Corticosteroids are controversial in the management of SJS/TEN |
| Aberer (2005) | Standardized approach to causality assessment | - Demoly questionnaire<br>- Lab-based: Skin testing, LTT, CAST-ELISA, 15-HETE-determination, flow cytometry basophil activation, leukocyte cytotoxic assay<br>- Provocation (challenge, re-challenge)<br>- Confirmatory testing: Prick and patch testing | - Literature review for drugs as complete/incomplete antigens<br>- In vivo algorithms for work-up of non-immediate hypersensitivity reactions<br>- Preparation of for patch testing<br>- Practice guidelines | - Most drugs are haptens not antigens, complicating testing because the drug-protein conjugate would need to be assessed, not the drug alone<br>- Skin prick test for IgE mediated and patch testing for delayed hypersensitivity<br>- Standardization of lab-based or confirmatory testing is not well established and has significant risks |
| Kotrulja (1997) | Diagnostic methods for drug allergy | - Skin testing: Prick, scratch, intradermal and conjunctival, patch, scratch-patch and photo-patch<br>- In vitro testing: RIST, RAST, ITDBG, LTT, CAST-ELISA<br>- Provocation: Oral challenge | - 17 different types of drug reactions<br>- Categorized based on Gell and Coombs types of hypersensitivity reaction | - Many drugs can interfere with the reaction to allergy skin testing (anti-histamines, TCA, chlorpromazine and benzodiazepines – type 1 HS; corticosteroids - type 4 HS.)<br>- RIST, RAST, ITBDG – type 1 HS (conduct 3-6 weeks after resolution of initial reaction)<br>- Use $\geq 4$ types of diagnostic testing to diagnose a drug allergy<br>- Consider oral challenge for conflicting results - keeping in mind risk benefit |
| Dubey (2006) | Reviews methods for causality assessment | - Clinical criteria helpful in defining CADR:<br>- 1) Exclude other causes (viral exanthema)<br>- 2) Temporal relationship (drug use and onset)<br>- 3) improvement following drug cessation<br>- 4) Reactivation upon re-challenge<br>- 5) Known drug/cutaneous reaction<br>- Skin biopsy, and estimation of drug levels in blood<br>- Oral re-exposure, or prick or scratch tests, may be carried out, after hospitalization | - Pathogenesis<br>- Risk factors<br>- Types of ADRs<br>- Clinical manifestations<br>- Diagnosis<br>- Differential diagnosis<br>- Management<br>- Preventative strategies | - Certain drug classes are most often implicated (ex. antibiotics and anticonvulsants)<br>- Symptomatic treatment may be accompanied by local skin care and, if indicated, immunomodulating therapy with corticosteroids to reduce the severity of the skin reaction<br>- If therapy with the offending drug is essential, it may be continued or reintroduced using previously published protocols |
| Ardern-Jones (2019) | Review and algorithm for complex cases | - Recent guidelines on optimal supportive care<br>- Interventional treatment for SJS/TEN<br>- Consider in-vitro diagnostics, HLA testing and skin testing before drug challenge testing | - Initial investigations<br>- Management of AGEP, DRESS, SJS/TEN<br>- Diagnostic testing | - Diagnostic testing is complicated by the potentially fatal outcome of drug re-exposure and the lack of consensus on the optimal acute therapy if a severe reaction is elicited |
| Sharma (1996) | Review of stepwise approach to suspected drug reactions | - In vivo testing: Rechallenge, patch testing, dechallenge<br>- In vitro testing: Radioallergosorbent test (RAST), ELISA, LTT, MIF, LTA, Basophil Degranulation Test, Histamine Release Test, Haemagglutination Assays 108 and flow cytometry | - Incidence<br>- Classification and mechanism<br>- Clinical manifestations<br>- Drugs associated with SJS/TEN<br>- Drug reactions in children and adolescents<br>- Stepwise approach: (1) History and physical examination, (2) In vivo testing, (3) In vitro testing. | - Most in vitro tests are unreliable for routine clinical use and are suitable only for immunologic research<br>- Short steroid courses have been advocated in severe maculopapular reactions, EM and SJS especially early in the disease (within 48 hours of onset) |
| Barbaud (2001) | Guidelines for skin tests in the investigation of CADRs | - Drug patch testing<br>- Prick tests<br>- Intradermal tests | - Guidelines in skin testing<br>- Drug imputability<br>- Guidelines in drug skin testing | - Success of skin tests varies with drug tested and features of the CADR<br>- If negative, a drug skin test does not exclude the responsibility of a drug<br>- If positive, determine its specificity and relevance<br>- Compare all skin test results with those obtained in negative controls, namely patients who have taken the drug within the last 6 months without adverse effects, or even patients who have never had contact with the culprit drug |
| Barbaud (2003) | Usefulness of patch testing in drug eruption | - Drug patch testing | - Analysis of guidelines for performing prick, ID and patch tests<br>- How to perform/interpret drug patch tests<br>- Drugs reported to yield positive reactions<br>- How to define the relevance and the specificity of drug patch tests in the workup of cutaneous drug allergies | - Value of drug patch tests depends on the clinical features of the CADR, the drug involved, the vehicle, the concentration used, and the site where they have been performed |
| Paulmann (2015) | Timelines, drug chart, classifying type of rash correctly and reviews in vitro | - EuroSCAR | - Clinical features<br>- Histology<br>- Differential diagnosis<br>- Epidemiology<br>- Etiology<br>- Therapy<br>- SCORTEN<br>- Allergy testing | - Severe drug reactions partly differ in the triggering agents and latency periods between intake and onset of initial symptoms<br>- Prognosis: SJS/TEN is poor and depends on patient's age, underlying conditions, extent of skin detachment; GBFDE is better but recurrences may lead to more severe disease manifestations; protracted and recurrent courses have been described in DRESS; AGEP resolves without problems |
| Bergmann (2019) | Role of in vivo and in vitro tests in the diagnosis of SCAR | - In vivo: Patch tests, delayed-reading skin prick tests, delayed-reading intradermal tests<br>- In vitro: Lymphocyte transformation test, LTT combined with cytokines and cytotoxic markers (cyto-LTT), cytokine determination by means of ELISA, ELISpot/bead assay, HLA-markers | - Sensitivity, advantages, and limitations of in vivo and in vitro tests in SCAR<br>- Proposed algorithm for culprit drug assessment with in vivo and in vitro tests in SCAR | - Patch testing remains first-line in most SCAR patients followed by delayed-reading IDTs in the case of negative results (note: IDTs are contraindicated in bullous diseases)<br>- In vitro tests have shown promising results in the diagnosis of SCAR. Positivity is high with cyto-LTT but this has to be confirmed with larger studies |
| Cabanas (2020) | Spanish guidelines for diagnosis, management, treatment and prevention of DRESS/DIHS syndrome | - Algorithm of the Spanish Pharmacovigilance System<br>- Patch tests<br>- Skin tests<br>- Lymphocyte transformation test | - Guidelines on the clinical and allergy diagnosis, management treatment and prevention of DRESS syndrome<br>- Practical recommendations (ASPS) | - Calculate the index day and perform causality assessment according to the ASPS as soon as DRESS diagnosis is suspected (at least score "Possible" using RegiSCAR)<br>- *In vitro* tests should not be performed before a minimal time interval of 4-8 weeks after the reaction and at least 4 weeks after stopping treatment with systemic corticosteroids<br>- LTT is the best documented assay for in vitro DRESS diagnosis<br>- Patch tests should be preceded by LTT or performed as first line in investigation of DRESS causality if in vitro tests are unavailable |
| Ingen-Housz-Oro. (2018) | National French diagnosis and care protocol for EN | - French guidelines for EN management | - ALDEN for SJS/TEN, drug chart and timeline | - Prompt withdrawal of culprit drug(s), transfer of the patient to a specialized unit, and maximal supportive care are the cornerstones of patient management |
| Cacoub (2011) | Literature review of DRESS syndrome | - Systematic review of retrospective case series DRESS | - Individual methods not studied | - DRESS should be highly suspected with the presence of skin rash, liver involvement, fever, hypereosinophilia, and lymphadenopathy<br>- High rate of HHV-6 and other herpes viruses reactivation associated with DRESS implies that HHV-6 and other herpes viruses should be detected in routine clinical practice |
| Ralph Edwards (2017) | Causality assessment in pharmacovigilance | - Algorithms: WHO, UMC<br>- Probabilistic | - Secondary effects<br>- Causal chain: Temporality, consistency, strength, specificity, dose-response, experimental support, plausible mechanism for effect, coherence, analogy | - Although we are not sure what level of risk is acceptable by the public at large, let alone those rare individuals that may have specific risk factors, we need to know much more about the type and incidence of all risks |
| Pande (2018) | Reviews commonly used CATs and its implications in clinical practice | - Clinical judgment<br>- Algorithmic methods: Naranjo, Kramer, Jones, Karch, Bégaud, Adverse Drug Reactions Advisory Committee guidelines<br>- Bayesian methods or probabilistic methods: BARDI | - Principles of causality assessment<br>- Challenges in developing causality assessment methods<br>- Methods of causality assessment | - There are multiple criteria or algorithms available in ADR, indicating that none of them is specific or complete<br>- Most of these causality assessment tools (CATs) use four cardinal principles of diagnosis: Temporal relationship of drug and drug reaction, biological plausibility of the drug causing a reaction, dechallenge, and rechallenge |
| Khan (2016) | Reviews Naranjo, Karch, Kramer, Bégaud, WHO-UMC, Liverpool, expert judgment | - Examines all drug causality tools (no focus on skin)<br>- Expert judgment (global inspection)<br>- Probabilistic method (logistic method)<br>- Algorithm methods<br>- Recent advances in causality assessment tools: Genetic, Liverpool, and Pediatric algorithms | - Scrutinize commonly used and recently developed methods of causality assessment tools for the diagnosis of adverse drug reactions and discuss their pros and cons<br>- Suggests a method universally applicable to all ADEs | - Rechallenge is frequently detrimental and impracticable<br>- Probabilistic methods are dependent on the preceding likelihood approximates that may not be there<br>- The current diagnosis of ADRs is empirically based, expert judgment<br>- Probabilistic methods are liable to generate cloudy causation results |
| Agabiaka (2008) | A systematic review of CAM for adverse drug reactions | - WHO & UMC, Naranjo<br>- BARDI<br>- Probabilistic<br>- Drug Interaction Probability Scale (DIPS)<br>- Karch and Lasagna, Kramer et al., Blanc et al., Emanuelli and Sacchetti, Evreux et al., Kitaguchi et al., Jones algorithm,<br>- French, Ciba-Geigy, and Australian methods | - Expert Judgement or global introspection<br>- Algorithms<br>- Probabilistic or Bayesian approaches<br>- Advantages and limitations | - The differences in ADR causality criteria and subjectivity of judgements may be responsible for the lack of reproducibility of most published methods<br>- No ADR causality assessment method has shown consistent measurement of causality; therefore, no single method is universally accepted |
| Meyboom (1997) | Review of the role of causality assessment in pharmacovigilance | - WHO, Naranjo, UMC<br>- Bayesian Adverse Reactions Diagnostic Instrument (BARDI)<br>- European ABO system<br>- Yale algorithm<br>- Karch and Lasagna<br>- Hallas et al. | - Efficiency<br>- Use of causality assessment in pharmacovigilance: Signal detection, regulatory measures, patient care<br>- Limitations<br>- Probability calculation<br>- Aetiological-diagnostic systems | - Causality assessment cannot eliminate uncertainty but can categorise it in a semiquantitative way<br>- Since none of the available systems has been validated, general case-causality assessment is of limited scientific value<br>- The causality categories developed by WHO have the advantage of being easy to use and internationally accepted |
| Meyboom (1998) | Review of causality assessment | - Naranjo<br>- Uppsala Monitoring Centre | - What standardized causality assessment can and cannot do<br>- Weighing the balance of evidence: Number of case reports, consistency of the data, time and dose relationship, plausibility of the hypothesis, experimental findings, analogies in previous experiences with other drugs, nature and quality of the data | Questions to address the role of causality assessment in pharmacovigilance:<br>- Should causality assessment only be done in selected case reports (e.g. unknown or serious adverse reactions)?<br>- Should one general system be used for all reactions or are special systems needed for specific adverse reactions? |

The benchmark, or standard, is a method assumed to be correct and can be used to assess or validate another CAM. Expert judgment was the most common standard used to validate other CAMs. Tissue biopsy was a component of the work-up in 27 studies (24.8%), and provocation/confirmatory testing (i.e. patch testing, prick testing, intradermal testing, challenge, re-challenge) was performed in 21 studies (19.3%). To statistically compare agreement between the benchmark method and the method(s) being studied, kappa values of agreement were sometimes included in studies.

Eleven publications (10.1%) studied lab-based methods for CDI either in the acute setting during a drug rash, or as confirmatory testing after an eruption had resolved. They can be divided into cytokine/cytotoxic-based testing and cell-based testing.

### Review article analysis

There were 26 review articles included. Each review article dealt with the management of ADEs (Table 7). A minority were explicitly skin focused and reviewed CAMs with one type of drug eruption or delineated methods used to assess, diagnose, and categorize CDEs. There were 14 reviews that studied lab-based methods and provocation testing. The latest review of CDI methods was in 2016 and discussed a select number of methods, their strengths, and weaknesses. In-depth comparisons between methods were not conducted. (8) Only 3 reviews quantitatively assessed CAMs by calculating sensitivity (SN), specificity (SP), positive predictive value (PPV) and negative predictive value (NPV).(6,12,13) The studies were heterogeneous and often did not assess CDEs separately from other types of ADEs. If a study utilized different scoring categories (e.g. 5 factor vs. 3 factor Likert scale) then they were often excluded from the review; therefore, not all published CAMs were actually studied.

### Diagnosing the type of drug eruption

Thirty-three types of CDEs were identified and the clinical and histologic characteristics, scoring systems, and commonly implicated drugs were investigated. SCARs have a significant morbidity and mortality risk. Clinical signs suggestive of SCARs include fever, prodromal malaise and fatigue, rapidly progressing rash, bullae/desquamation, Nikolsky sign, mucosal involvement, and systemic findings (liver, kidney, thyroid, GI, cardiac abnormalities). Risk factors have been identified that are associated with increased frequency and severity of drug hypersensitivity reactions. Specific risk factors are discussed in a subsequent section.

### Latency between drug exposure and onset of rash

The latencies between drug exposure and rash onset are known to vary depending on drug rash type. Over 75% of published CAMs utilized time from drug exposure to rash onset as a criterion. It was also observed that there were drug class specific latencies, suggesting that drug rash type is not the only factor affecting latency. For example, a relatively shorter exposure to rash latency was seen with radio contrast dye (<6h) as well as antibiotics and acetaminophen/NSAIDs (<15 days). Drugs found to have longer latencies include anticonvulsants (>15 days) and xanthine oxidase inhibitors (21-90 days). (14,15)

These trends were noted when controlling for drug rash type in some publications, but more robust, larger studies are needed to confirm these associations. (16–18) Anticonvulsant hypersensitivity had a mean latency of 28 days when causing DRESS, an eruption known to have a longer latency. When DRESS was caused by an antibiotic, a drug class shown to have a shorter latency, the average delay was much shorter (∼18 days). (19) Another factor affecting latency occurs with re-exposure to the causative drug. It is well established that re-exposure (re-challenge) to a culprit drug shortens the latency independent of rash type or medication class. (20) While absolute latency ranges were very broad, most cases of a certain eruption type had either long or shot latencies on average. For example, eruptions with longer latencies include DRESS (∼14-42 days) and SJS/TEN (∼7-21 days), and eruptions with shorter latencies include urticaria (<24h), morbilliform drug eruption (∼4-14 days), AGEP (<4 days), and fixed drug eruption (∼7-14 days). (1,17,21)

If latencies differ by drug rash type, or drug class, it is possible that erroneously narrowing the latency range may result in a false negative causality assessment, missing the culprit drug and stopping the wrong medication.

### Risk factors and associated conditions

Risk factors for severity and frequency of CDEs have been identified including: Polypharmacy, old age, female sex, re-exposure, liver and kidney disease, diabetes, HIV, malignancy, and certain HLA subtypes (14,19,22,23). Many of these risk factors are integrated into select diagnostic and prognostication tools (e.g DRESS RegiSCAR criteria, SCORTEN for SJS/TEN, RegiSCAR AGEP score). (14,19,22,23) Co-morbidities were also seen as important for ruling out non-drug related conditions (e.g. auto-immune disease, and infection). Accurate characterization of these risk factors as they relate to drug eruptions, and their prognostic significance should be considered when managing CDEs.

### Causality methods comparison

This review identified 54 published CAMs. They share many similarities, differences, strengths, and limitations. CAMs were first compared based on their respective components (Table 5). Algorithmic methods tended to have more questions related to clinical changes with exposure to the suspected drug (challenge), discontinuation (de-challenge) and re-exposure (re-challenge). Probabilistic approaches tended to have more criteria related to ruling out other causes of the reaction, assessment of other potential culprit medications and determining how often the suspected drug had been reported to cause the CDE. Drug exposure to rash latency was a criterion in 66.0% of published methods. Prior sensitization was a criterion for 34.6% of methods (Algorithmic 45.5%, Probabilistic 16.7%). Considering an alternate diagnosis/cause was a criterion in 57.4% (algorithmic 67.7%, probabilistic 38.9%). An alternate drug cause or drug notoriety was a criterion in 31.5% (algorithmic 20.6%, probabilistic 44.4%). Considering dose changes and therapeutic drug levels were part of 44.2% of methods (algorithmic 60.6%, probabilistic 11.1%). Challenge, de-challenge, and rechallenge were criteria for 20.4%, 48.1, 60.4%, respectively (algorithm: 26.5%, 58.8%, 76.5%; Probabilistic: 5.6%, 22.2% and 33.3%). Causality assessment ratings were often reported on scales of varying levels such as the 5-point Likert scale (e.g. Very likely, likely, possible, unlikely, very unlikely). A 5-point scale for probability of causality was found in 25.9% of methods, of which 78.6% were algorithms, 7.1% probabilistic, and the remaining 14.3% from expert judgment CAMs. Less than a 5-point scale for probability of causality was found in 74.1% of methods, of which 57.5% were algorithms and 42.5% probabilistic approaches.

A number of studies and several review articles reported the efficacy (i.e. SN, PPV, SP, NPV) of many published methods to display how studies have attempted to validate, compare, and analyze the CAMs (See Table 6). Generally, algorithms had relatively higher SN and PPV, but lower SP and NPV. Probabilistic approaches generally had lower SN and PPV, but higher SP and NPV. Methods that combine elements of the different categories were found to be more effective in several studies. (24–26) Additional studies, such as with a systematic review, are required to determine the significance of this observation. Expert judgment was the most common CAM used as the standard when validating other methods, but was repeatedly noted to be subjective, have lower reproducibility and inter/intra-rater agreement.

There were 9 lab-based culprit drug investigations published in 11 studies and 14 reviews. They were mainly based on measurement of immune cells or inflammatory cytokine/chemokine parameters. The main tests included lymphocyte transformation test (LTT), interferon gamma release assay, CellScan technique, histamine releasing test (HRT), HLA-B allelic variation, granzyme B-ELISpot assay, granulysin expression, cytokine beads array assay, and chemi-informatics based QSAR model (quantitative structure-activity relationship).

## DISCUSSION

### Summary of evidence

Drug reactions pose a significant health problem, and are costly to the healthcare system. (27) ADEs are heterogeneous, and often affect more than one organ system. The most common organ for early manifestation of ADEs is the skin. Cutaneous manifestations may allow for a more accurate timeline compared to internal involvement which can often be sub-clinical. This may translate to more efficacious causality assessment. Developing CAMs using ADE databases may not always be accurate because of the heterogeneity between drug exposure, cutaneous and systemic manifestations. CAMs designed for CDE may, therefore, be higher yield, and easier to validate.

When dealing with drug eruptions, important aspects of the patient history include degree/duration of drug exposure, time of rash onset, confounding factors (e.g. comorbidities), rechallenge (re-exposure to drug), dechallenge (improvement with removal of the causative drug), and background epidemiological and clinical information. This data is also vital for accurate pharmacovigilance adverse event reporting (6)

### Medication history and onset of cutaneous eruption

Accurate documentation regarding medication administration (i.e. when drug was started, last taken, and dose increases) is important in order to determine the latency between possible drug exposures and the reaction. CDE may allow for the most accurate determination of latency as cutaneous manifestations are often an early finding compared to signs of end organ damage. Studying CAMs for CDE separate from non-skin ADE could potentially ameliorate the issue of heterogenicity and problematic validation methods. (2,20,28)

### Determining the type of eruption and causative drug

Different types of drug eruptions have different causative drugs and different latencies between exposure and rash onset. This review has demonstrated that in addition to drug rash type, drug classes may also play an important role in affecting this latency. If drug class specific latency ranges can be standardized in conjunction with rash type, the latency window could be narrowed, increasing accuracy and decreasing confounding by concurrent medications and conditions. As we learn more about the pathophysiology of different drug rashes, we may uncover potential diagnostic and therapeutic markers using lab-based techniques, but the importance of clinical methods must be underscored, especially with rapidly progressive and deadly SCARs. (29)

Tissue biopsy can often be helpful to determine the type of drug eruption or rule out non-drug related conditions, but often have non-specific findings and may delay appropriate management.

### Causality assessment methods

A systematic review of causality assessment with adverse drug reactions in 2008, identified approximately 40 different CAMs, but did not specifically address the issue of cutaneous adverse drug events. (6) The information gathered was very heterogeneous and effect modification may have obscured certain findings because of the inherent differences between clinical behaviour and diagnostic accuracy with cutaneous versus other types of ADEs. The current review has identified 54 CAMs, not including investigative or lab-based techniques. The number of published methods continues to grow, yet the data suggests we are no closer to being able to reliably test the efficacy of any proposed methods, nor develop guidelines to guide causality assessment. The findings also show that compared to systemic/internal ADEs, cutaneous adverse reactions may be easier to diagnose early. There are also more accessible investigations available to diagnose CDEs (e.g. morphology and evolution, tissue biopsy, direct immunofluorescence, indirect immunofluorescence, in vitro blood tests, skin based confirmatory testing and HLA-typing). This may allow for more accurate ascertainment of date of onset, progression, and improvement over time, making the study of CAMS more effective and may even serve as the benchmark to appraise and develop consensus guidelines for CDI in the future.

There are three main categories of ‘causality assessment and most of the proposed methods fall into one or more of them. Algorithmic approaches (i.e. operational algorithms), were found to be the most commonly studied method, had high sensitivity and PPV, and were easiest to use. Probabilistic approaches were the second most commonly studied method and had the highest specificity and NPV, but were time consuming, complex, and not always practical. Lastly, expert judgment, which was often used as the standard when studying other methods, was subjective, and had poor intra/inter-rater reproducibility. Confounding variables, including multiple medications and co-morbidities, compromise algorithmic sensitivity and specificity. (6)

A small number of studies compared more than one CAM and when they studied a method that used aspects of more than one category (e.g. combined algorithmic and probabilistic approaches), they were found to have higher efficacy and reduced some limitations seen in the methods used separately. (3,26,30) It has been suggested that greater accuracy may be obtained by first employing the method with the highest sensitivity to capture true positive cases (e.g. algorithmic approach), followed by employing a method with greater specificity to rule out the greatest number of true negatives (e.g. probabilistic method), resulting in a higher percentage of true positives than either method alone.

### Investigations: Lab-based, pharmacogenomic and confirmatory testing

Lab-based investigations are becoming more prevalent and may complement clinical CAMs. We identified 11 studies and 14 reviews that compared various types of in vitro testing: lymphocyte transformation test (LTT), interferon gamma release assay, CellScan technique, histamine releasing test (HRT), HLA-B allelic variation, granzyme B-ELISpot assay, granulysin expression, cytokine beads array assay, and chemi-informatics based QSAR model (quantitative structure-activity relationship). Limitations of lab-based studies included cost, expertise needed to conduct tests, interpretation of results, and the fact that lab-based ex vivo investigations may not be representative of in vivo conditions. As well, allergies to drug intermediates or drug-carrier protein conjugates may result in false negatives because the techniques often test the parent drug and not the intermediate or metabolite that may actually be the culprit such as with acrolein, the metabolite of cyclophosphamide, responsible for hemorrhagic cystitis, a serious ADR. (31,32) Generally, they have shown promising results, but with significant variability. If sufficiently validated, these investigations may be useful alternatives to the riskier current gold standard, which is re-exposure to the culprit drug.

Methods of confirmatory testing include re-exposure or provocation testing which can be done through oral or cutaneous modalities such as with patch, prick and intradermal testing. The efficacy of this type of testing varies on drug eruption type and causative drug. There is also a risk for severe reactions caused by re-exposure to the culprit drug, even in small quantities. Confirmatory/provocation testing usually needs to be performed 4-6 weeks after the eruption has resolved. Patch testing was reported to be most accurate for SDRIFE, AGEP, FDE, DRESS, MDE, and to a lesser degree SJS/TEN. Cross-reactivity between different antiepileptic drugs was also detectable. (33–36).

### Limitations

This scoping review intended to collate all published, peer-reviewed works relating to CDI. Limitations include the possibility that our search strategy did not to capture all appropriate papers. This was mitigated by scanning the references of included papers to identify sources not captured by our search strategy. A scoping review is descriptive and meant to gather available information rather than attempt to statistically compare published methods, conduct meta-analyses, or determine superiority. Cutaneous reactions may be easier to identify by patients and physicians than systemic ADEs. Some of the included studies used national and international ADE registries that did not always specify if the ADE was cutaneous or systemic. Some of the CAMs were studied using these databases containing all types of drug reactions. This may have contributed to why some of the studied CAMs performed poorly and with low reproducibility. Standard, or benchmark, methods for validating or comparing CAMs can be problematic. The accuracy of the standard, which was most commonly expert judgment or provocation testing, may themselves be inaccurate. Additionally, many CAMs do not account for polypharmacy, especially when drugs are initiated concomitantly. Stopping multiple drugs may leave other co-morbidities untreated which may be a potential confounder when assessing ADEs severity, response to treatment and resolution after stopping the culprit drug.

## CONCLUSIONS

The mainstay of managing drug eruptions is identifying and stopping the culprit drug, which can be challenging. This review has synthesized the highest quality published methods available, focusing on CDEs. While none of the 54 published methods have been shown to be the most effective, consistently accurate, or widely applicable, some have shown promise, including CAMs that combine more than one category (e.g. operational algorithms combined with probabilistic approaches), lab-based methods to test for drug specific immune cells, and HLA-based pharmacogenomic risk assessment. Confirmatory testing such as with provocation/re-challenge, patch testing, and intradermal testing are currently the gold standard for correct drug identification, but they too can have variable efficacy, risks, and are mainly applicable 4-6 weeks after the eruption has subsided. From this scoping review, more studies can be conducted that integrate the strengths of published CAMs and avoid the limitations seen in past methods.

## Supporting information

PRISMA-ScR Guidelines

## Data Availability

Sources of evidence: Medline, Embase, and Cochrane Central Register of Controlled Trials.

## ABBREVIATIONS AND ACRONYMS

ADE: Adverse drug event
AGEP: Acute generalized exanthematous pustulosis
CAM: Causality assessment method
CDE: Cutaneous drug eruption
CDI: Culprit drug identification
DIHS: Drug induced hypersensitivity syndrome
DRESS: Drug reaction with eosinophilia and systemic signs
EM: Erythema multiforme
FDE: Fixed drug eruption
HHV6 & 7: Human herpes virus 6 & 7
HRT: Histamine release assay
LTT: Lymphocyte transformation test
NPV: Negative predictive value
PPV: Positive predictive value
SJS: Stevens Jonson syndrome
SN: Sensitivity
SP: Specificity
TEN: Toxic epidermal necrolysis

**Appendix**

## References

1. Mockenhaupt M. Epidemiology of cutaneous adverse drug reactions. Advers Cutan Drug Eruptions. 2017;1(1):96–108.

2. Demoly, P; Adkinson, N. F; Brockow, K; Castells, M; Chiriac, A. M; Greenberger, P. A; Khan, D. A; Lang, D. M; Park, H.-S; Pichler, W; Sanchez-Borges, M; Shiohara, T; Thong BY-H. International consensus on drug allergy. Allergy Eur J Allergy Clin Immunol. 2014;69(4):420–37.

3. Théophile H, André M, Arimone Y, Haramburu F, Miremont-Salamé G, Bégaud B. An updated method improved the assessment of adverse drug reaction in routine pharmacovigilance. J Clin Epidemiol. 2012;

4. Duong Tu Anh; Valeyrie-Allanore, Laurence; Wolkenstein, Pierre; Chosidow O. Severe cutaneous adverse reactions to drugs. Lancet. 2017;390(10106):1996–2011.

5. Théophile H, Dutertre JP, Gérardin M, Valnet-Rabier MB, Bidault I, Guy C, et al. Validation and reproducibility of the updated French causality assessment method: An evaluation by pharmacovigilance centres & pharmaceutical companies. Therapie. 2015;70(5):465–76.

6. Agbabiaka Taofikat B; Savovic Jelena Ernst E. Methods for causality assessment of adverse drug reactions: A systematic review. Drug Saf. 2008;31(1):21–37.

7. Théophile H, André M, Miremont-Salamé G, Arimone Y, Bégaud B. Comparison of three methods (an updated logistic probabilistic method, the Naranjo and Liverpool algorithms) for the evaluation of routine pharmacovigilance case reports using consensual expert judgement as reference. Drug Saf. 2012;65(10):1069–77.

8. Khan LM, Al-Harthi SE, Osman AMM, Sattar MAAA, Ali AS. Dilemmas of the causality assessment tools in the diagnosis of adverse drug reactions. Saudi Pharm J. 2016;24(4):485–93.

9. Danan G, Benichou C. Causality assessment of adverse reactions to drugs-I. A novel method based on the conclusions of international consensus meetings: Application to drug-induced liver injuries. J Clin Epidemiol. 1993;46(11):1323–30.

10. Arimone, Yannick; Bégaud, Bernard; Miremont-Salamé, Ghada; Fourrier-Réglat, Annie; Molimard, Mathieu; Moore, Nicholas; Haramburu F. A new method for assessing drug causation provided agreement with experts’ judgment. J Clin Epidemiol. 2006;59(3):308–14.

11. Tricco Andrea C; Lillie, Erin; Zarin, Wasifa; O’Brien, Kelly K; Colquhoun, Heather; Levac, Danielle; Moher, David; Peters, Micah D.J; Horsley, Tanya; Weeks, Laura; Hempel, Susanne; Akl, Elie A; Chang, Christine; McGowan, Jessie; Stewart, Lesley; Hartling SE. PRISMA extension for scoping reviews (PRISMA-ScR): Checklist and explanation. Ann Intern Med. 2018;169(7):467.

12. Daoud MS, Schanbacher CF, Dicken CH. Recognizing cutaneous drug eruptions. Reaction patterns provide clues to causes. Postgrad Med. 1998;104(1):101–5.

13. Macedo AF, Marques FB, Ribeiro CF, Teixeira F. Causality assessment of adverse drug reactions: Comparison of the results obtained from published decisional algorithms and from the evaluations of an expert panel. Pharmacoepidemiol Drug Saf. 2005;14(12):885–90.

14. Atzori L., Pinna A. L., Pilloni L., Ferreli C., Aste N., Zucca M., Pau M. AN. Acute generalized exanthematous pustulosis: the experience of an Italian drug-surveillance centre. G Ital di Dermatologia e Venereol. 2007;142(4):303–10.

15. Davidovici, Batya; Dodiuk-Gad, Roni; Rozenman, Dganit; Halevy S. Profile of acute generalized exanthematous pustulosis in Israel during 2002-2005: results of the RegiSCAR Study. Isr Med Assoc J. 2008;10(6):410–2.

16. Akpinar, Fatma; Dervis E. Drug eruptions: An 8-year study including 106 inpatients at a dermatology clinic in Turkey. Indian J Dermatol. 2012;57(3):194–8.

17. Soria A, Bernier C, Veyrac G, Barbaud A, Puymirat E, Milpied B. Drug reaction with eosinophilia and systemic symptoms may occur within 2 weeks of drug exposure: A retrospective study. J Am Acad Dermatol. 2020;82(3):606–11.

18. Scavone C, Di Mauro C, Ruggiero R, Bernardi FF, Trama U, Aiezza ML, et al. Severe cutaneous adverse drug reactions associated with allopurinol: An analysis of spontaneous reporting system in Southern Italy. Drugs - Real World Outcomes. 2020;7(1):41–51.

19. Kardaun S, Sekula P, Valeyrie-Allanore L, Liss Y, Chu C, Creamer D, et al. Drug reaction with eosinophilia and systemic symptoms (DRESS): An original multisystem adverse drug reaction. Results from the prospective RegiSCAR study. Br J Dermatol. 2013;169(5):1071–80.

20. Brockow K, Przybilla B, Aberer W, Bircher AJ, Brehler R, Dickel H, et al. Guideline for the diagnosis of drug hypersensitivity reactions. Allergo J Int. 2015;

21. Zhang C, Van DN, Hieu C, Craig T. Drug-induced severe cutaneous adverse reactions: Determine the cause and prevention. Ann Allergy Asthma Immunol. 2019;123(5 PG-483–487):483–7.

22. Alniemi Dema T; Wetter David A; Bridges, Alina G; el‐Azhary, Rokea A; Davis, Mark D. P; Camilleri, Michael J; McEvoy MT. Acute generalized exanthematous pustulosis: clinical characteristics, etiologic associations, treatments, and outcomes in a series of 28 patients at Mayo Clinic, 1996-2013. Int J Dermatol. 2017;56(4):405–14.

23. Bastuji-Garin S, Fouchard N, Bertocchi M, Roujeau JC, Revuz J, Wolkenstein P. Scorten: A severity-of-illness score for toxic epidermal necrolysis. J Invest Dermatol. 2000;115(2):149–53.

24. Théophile H, Arimone Y, Miremont-Salamé G, Moore N, Fourrier-Réglat A, Haramburu F, et al. Comparison of three methods (Consensual expert judgement, algorithmic and probabilistic approaches) of causality assessment of adverse drug reactions: An assessment using reports made to a French pharmacovigilance centre. Drug Saf. 2010;33(11):1045–54.

25. Théophile, Hélène; André, Manon; Miremont-Salamé, Ghada; Arimone, Yannick; Bégaud B. c. Drug Saf. 2013;36(10):1033–44.

26. Zhao J, Hu L, Zhang L, Zhou M, Gao L, Cheng L, et al. Comparison of three methods (an updated logistic probabilistic method, the Naranjo and Liverpool algorithms) for the evaluation of routine pharmacovigilance case reports using consensual expert judgement as reference. Drug Saf. 2017;10(10):194–8.

27. Palanisamy Sivanandy, Kumaran KoSA RA. A study on assessment, monitoring and reporting of adverse drug reactions in Indian hospital. Asian J Pharm Clin Res. 2011;4(3):112–6.

28. Benahmed S, Picot MC, Hillaire-Buys D, Blayac JP, Dujols P, Demoly P. Comparison of pharmacovigilance algorithms in drug hypersensitivity reactions. Eur J Clin Pharmacol. 2005;61(7):537–41.

29. Chave, T.A; Mortimer, N.J; Sladden, M.J; Hall, A.P; Hutchinson P. Toxic epidermal necrolysis:current evidence, practical management and future directions. Br J Dermatol. 2005;153(2):241–53.

30. Naranjo CA, Kwok MC, Lanctôt KL, Zhao H-P, Spielberg SP, Shear NH. Enhanced differential diagnosis of anticonvulsant hypersensitivity reactions by an integrated Bayesian and biochemical approach. Clin Pharmacol Ther. 1994;56(5):564–75.

31. Salameh, Fares; Kravitz, Martine Szyper; Barzilai, Aviv; Baum, Sharon; Shoenfeld, Yehuda; Schiffenbauer, Yael; Trubniykov, Ela; Trau H. Clinical application of static fluorescence-based cytometry, Cellscan, in cutaneous adverse drug reaction. J Dermatol. 2011;38(5):447–55.

32. Low Yen S; Caster, Ola; Bergvall, Tomas; Fourches, Denis; Zang, Xiaoling; Norén, G Niklas; Rusyn, Ivan; Edwards, Ralph; Tropsha A. Cheminformatics-aided pharmacovigilance: Application to Stevens-Johnson Syndrome. J Am Med Informatics Assoc. 2016;23(5):968–78.

33. Zeghal LBMNBHGBKAHZSSKK. Epicutaneous patch testing in delayed drug hypersensitivity reactions induced by antiepileptic drugs. Therapie. 2017;72(5):539–45.

34. Santiago, Felicidade; Gonçalo, Margarida; Vieira, Ricardo; Coelho Sónia; Figueiredo A. Epicutaneous patch testing in drug hypersensitivity syndrome (DRESS). Contact Dermatitis. 2010;62(1):47–53.

35. Friedmann Peter S; Ardern-Jones M. Patch testing in drug allergy. Curr Opin Allergy Clin Immunol. 2010;10(4):291–6.

36. Barbaud A. Usefulness of drug patch testing in cutaneous drug allergy: What is new? Rev Fr d’allergologie d’immunologie Clin. 2003;43(4):222–6.

37. Aceves-Avila, Francisco Javier; Benites-Godínez V. Drug allergies may be more frequent in systemic lupus erythematosus than in rheumatoid arthritis. J Clin Rheumatol. 2008;14(5):261–3.

38. Atzori L., Pinna A. L. Ferreli C. AN. Adverse cutaneous reactions to cardiovascular drugs: the experience of the department of dermatology in Cagliari. G Ital di Dermatologia e Venereol. 2006;141(2):123–30.

39. Barvaliya, M; Sanmukhani, J; Patel, T; Paliwal, N; Shah, H; Tripathi C. Drug-induced Stevens-Johnson syndrome (SJS), toxic epidermal necrolysis (TEN), and SJS-TEN overlap: A multicentric retrospective study. J Postgrad Med. 2011;57(2):115–9.

40. S Bastuji-Garin 1, M Zahedi, JC Guillaume JCR. Toxic epidermal necrolysis (Lyell syndrome) in 77 elderly patients. Age Ageing. 1993;22(6):450–6.

41. Beniwal, Ranjana;Gupta, Lalit; Khare, Ashok; Mittal, Asit; Mehta, Sharad; Balai M. Clinical profile and comparison of causality assessment tools in cutaneous adverse drug reactions. Indian Dermatol Online J. 2019;10(1):27–33.

42. Borch, JE; Andersen, KE; Bindslev-Jensen C. The prevalence of acute cutaneous drug reactions in a Scandinavian university hospital. Acta Derm Venereol. 2006;86(6):518–22.

43. Cabañas, R; Calderón, O; Ramírez, E; Fiandor, A; Caballero, T; Heredia, R; Herranz, P; Madero, R; Quirce, S; Bellón T. Sensitivity and specificity of the lymphocyte transformation test in drug reaction with eosinophilia and systemic symptoms causality assessment. Clin Exp allergy. 2018;48(3):325–33.

44. Chattopadhyay, C; Chakrabarti N. A cross-sectional study of cutaneous drug reactions in a private dental college and government medical college in eastern India. Niger J Clin Pract. 2012;15(2):194–8.

45. Das, Sudip; Roy, Aloke; Biswas I. A six-month prospective study to find out the treatment outcome, prognosis and offending drugs in toxic epidermal necrolysis from an urban institution in Kolkata. Indian J Dermatol. 2013;58(3):191–3.

46. Devi, K; George, Sandhya; Criton, S; Suja, V; Sridevi PK. Carbamazepine--the commonest cause of toxic epidermal necrolysis and Stevens-Johnson syndrome: a study of 7 years. Indian J Dermatol Venereol Leprol. 2005;71(5):325–8.

47. Drago, Francesco; Cogorno, Ludovica; Agnoletti, Arianna Fay; Ciccarese, Giulia; Parodi A. A retrospective study of cutaneous drug reactions in an outpatient population. Int J Clin Pharm. 2015;37(5):739–43.

48. East-Innis, AD; Thompson DS. Cutaneous drug reactions in patients admitted to the dermatology unit at the University Hospital of the West Indies, Kingston, Jamaica. West Indian Med J. 2009;58(3):227–30.

49. Eman A El-Nabarawy, Maha Fathy Elmasry, Mona Ibrahim El Lawindi YAT. An epidemiological and clinical analysis of cutaneous adverse drug reactions seen in a tertiary care outpatient clinic in Cairo, Egypt. Acta Dermatovenerologica Croat. 2018;26(3):233–42.

50. Frey, Noel; Bodmer, Michael; Bircher, Andreas; Jick, Susan S; Meier, Christoph R; Spoendlin J. Stevens-Johnson syndrome and toxic epidermal necrolysis in association with commonly prescribed drugs in outpatient care other than anti-epileptic drugs and antibiotics: A population-based case-control study. Drug Saf. 2019;42(1):55–66.

51. Frey, Noel; Bodmer, Michael; Bircher, Andreas; Rüegg, Stephan; Jick, Susan S; Meier, Christoph R; Spoendlin J. The risk of Stevens-Johnson syndrome and toxic epidermal necrolysis in new users of antiepileptic drugs. Epilepsia. 2017;58(12):2178–85.

52. Gholami, K; Parsa, S; Shalviri, G; Sharifzadeh, M; Assasi N. Anti-infectives-induced adverse drug reactions in hospitalized patients. Pharmacoepidemiol Drug Saf. 2005;14(7):501–6.

53. Acharya, LeelavathiD; Rao, PadmaGM; Ghosh S. Study and evaluation of the various cutaneous adverse drug reactions in Kasturba hospital, Manipal. Indian J Pharm Sci. 2006;68(2):212–5.

54. Goldberg, Ilan; Hanson, Meital; Chodick, Gabriel; Shirazi, Idit; Brenner, Sarah; Maggi E. In vitro release of interferon-gamma from peripheral blood lymphocytes in cutaneous adverse drug reactions. Clin Dev Immunol. 2012;2012:687532–7.

55. Goldberg, Ilan; Gilburd, Boris; Kravitz, Martine Szyper; Kivity, Shmuel; Chaim, Berta Ben; Klein, Tirza; Schiffenbauer, Yael; Trubniykovr, Ela; Brenner, Sarah; Shoenfeld Y. A novel system to diagnose cutaneous adverse drug reactions employing the cellscan--comparison with histamine releasing test and Inf-γ releasing test. Clin Dev Immunol. 2005;12(1):85–90.

56. Goldma Jennifer L; Chung, Wen-Hung; Lee, Brian R; Chen, Chun-Bing; Lu, Chun-Wei; Hoetzenecker, Wolfram; Micheletti, Robert; Yasuda, Sally Usdin; Margolis, David J; Shear, Neil H; Struewing, Jeffery P; Pirmohamed M. Adverse drug reaction causality assessment tools for drug-induced Stevens-Johnson syndrome and toxic epidermal necrolysis: room for improvement. Eur J Clin Pharmacol. 2019;75(8):1135–41.

57. Jennifer R Grace, Anna K Saina, Maheswari E, Srinivasa R VS. Assessment of adverse drug reactions occurring at department of neurology of a tertiary care hospital in India. Asian J Pharm Clin Res. 2018;11(10):457–64.

58. Hassoun-Kheir, Nasreen; Bergman, Reuven; Weltfriend S. The use of patch tests in the diagnosis of delayed hypersensitivity drug eruptions. Int J Dermatol. 2016;55(11):1219–24.

59. He Y, Chong FHT, Lim J, Lee RJT, Yap CW. Determination of the potential of drug candidates to cause severe skin disorders using computational modeling. Mol Inform. 2013;32(3):303–12.

60. Hirapara H, Patel T, Barvaliya M, Tripathi C. Drug-induced Stevens-Johnson syndrome in Indian population: A multicentric retrospective analysis. Niger J Clin Pract. 2017;20(8):978–83.

61. Sd I, M M, Ns M, Amutha A, I GJ, Rahman F. Pharmacovigilance of the cutaneous drug reactions in outpatients of dermatology department at a tertiary care hospital. J Clin Diagnostic Res. 2012;6(10):1688–91.

62. Jain S, Katiyar P, Suvirya S, Verma P, Sachan A, Nath R, et al. Study of therapeutic outcome and monitoring of adverse drug reactions (ADRs) in patients coming to outdoor patient department (OPD) of dermatology, venereology and leprosy in tertiary care hospital of Northern India. Int J Pharm Sci Res. 2020;11(1):474–88.

63. James J, Rani J. A prospective study of adverse drug reactions in a tertiary care hospital in South India. Asian J Pharm Clin Res. 2020;13(1):89–92.

64. Jung HY, Park S, Shin B, Lee J-H, Lee SJ, Lee MK, et al. Prevalence and clinical features of drug reactions with eosinophilia and systemic symptoms syndrome caused by antituberculosis drugs: A retrospective cohort study. Allergy Asthma Immunol Res. 2019;11(1):90–103.

65. Kano Y, Hirahara K, Mitsuyama Y, Takahashi R, Shiohara T. Utility of the lymphocyte transformation test in the diagnosis of drug sensitivity: Dependence on its timing and the type of drug eruption. Allergy. 2007;62(12):1439–44.

66. Karami Z, Mesdaghi M, Karimzadeh P, Mansouri M, Taghdiri MM, Kayhanidoost Z, et al. Evaluation of lymphocyte transformation test results in patients with delayed hypersensitivity reactions following the use of anticonvulsant drugs. Int Arch Allergy Immunol. 2016;170(3):158– 62.

67. Kato K, Kawase A, Azukizawa H, Hanafusa T, Nakagawa Y, Murota H, et al. Novel interferon-gamma enzyme-linked immunoSpot assay using activated cells for identifying hypersensitivity-inducing drug culprits. J Dermatol Sci. 2017;86(3):222–9.

68. Klaewsongkram J, Sukasem C, Thantiworasit P, Suthumchai N, Rerknimitr P, Tuchinda P, et al. Analysis of HLA-B allelic variation and IFN-gamma ELISpot responses in patients with severe cutaneous adverse reactions associated with drugs. J Allergy Clin Immunol Pract. 2019;7(1):219– 27.

69. Koelblinger P, Dabade TS, Gustafson CJ, Davis SA, Yentzer BA, Kiracofe EA, et al. Skin manifestations of outpatient adverse drug events in the United States: A national analysis. J Cutan Med Surg. 2013;17(4):269–75.

70. Kumar Das K, Khondokar S, Rahman A, Chakraborty A. Unidentified drugs in traditional medications causing toxic epidermal necrolysis: A developing country experience. Int J Dermatol. 2014;53(4):510–5.

71. Kurle D, Jalgaonkar S, Daberao V, Chikhalkar S, Raut S. Study of clinical and histopathological pattern, severity, causality and cost analysis in hospitalised patients with cutaneous adverse drug reactions in a tertiary care hospital. Int J Pharm Sci Res. 2018;9(5):1857–64.

72. Lange-Asschenfeldt C, Grohmann R, Lange-Asschenfeldt B, Engel RR, Ruther E, Cordes J. Cutaneous adverse reactions to psychotropic drugs: Data from a multicenter surveillance program. J Clin Psychiatry. 2009;70(9):1258–65.

73. Lebrun-Vignes B, Guy C,Jean-Pastor M-J, Gras-Champel V, Zenut M. Is acetaminophen associated with a risk of Stevens-Johnson syndrome and toxic epidermal necrolysis? Analysis of the French Pharmacovigilance Database. Br J Clin Pharmacol. 2018;84(2):331–8.

74. Lin M, Dai Y, Pwu R, Chen Y, Chang N. Risk estimates for drugs suspected of being associated with Stevens-Johnson syndrome and toxic epidermal necrolysis: A case-control study. Intern Med J. 2005;35(3):188–90.

75. Lin Y-F, Yang C-H, Sindy H, Lin J-Y, Rosaline Hui C-Y, Tsai Y-C, et al. Severe cutaneous adverse reactions related to systemic antibiotics. Clin Infect Dis. 2014;58(10):1377–85.

76. Loo CH, Tan WC, Khor YH, Chan LC. A 10-years retrospective study on Severe Cutaneous Adverse Reactions (SCARs) in a tertiary hospital in Penang, Malaysia. Med J Malaysia. 2018;73(2):73–7.

77. Maggio N, Firer M, Zaid H, Bederovsky Y, Aboukaoud M, Gandelman-Marton R, et al. Causative drugs of Stevens-Johnson syndrome and toxic epidermal necrolysis in Israel. J Clin Pharmacol. 2017;57(7):823–9.

78. Malhotra S, Chopra S, Gupta C, Dogra A. Cutaneous adverse drug reactions in a North Indian Tertiary Care Hospital. Int J Risk Saf Med. 2006;18(2):91–7.

79. Miliszewski MA, Kirchhof MG, Sikora S, Papp A, Dutz JP. Stevens-Johnson syndrome and toxic epidermal necrolysis: An analysis of triggers and implications for improving prevention. Am J Med. 2016;129(11):1221–5.

80. Mittal N, Gupta M, Singla M. Cutaneous adverse drug reactions notified by pharmacovigilance in a tertiary care hospital in north India. Cutan Ocul Toxicol. 2014;33(4):289–93.

81. Modi A, Desai M, Shah S, Shah B. Analysis of cutaneous adverse drug reactions reported at the Regional ADR Monitoring Center. Indian J Dermatol. 2019;64(3):250.

82. S P, K M, S A. Causality, severity and preventability assessment of adverse cutaneous drug reaction: A prospective observational study in a tertiary care hospital. J Clin Diagnostic Res. 2013;7(12):2765–7.

83. Papay J, Yuen N, Powell G, Mockenhaupt M, Bogenrieder T. Spontaneous adverse event reports of Stevens-Johnson syndrome/toxic epidermal necrolysis: Detecting associations with medications. Pharmacoepidemiol Drug Saf. 2012;21(3):289–96.

84. Park CS, Kang DY, Kang MG, Kim S, Ye YM, Kim SH, et al. Severe cutaneous adverse reactions to antiepileptic drugs: A nationwide registry-based study in Korea. Allergy Asthma Immunol Res. 2019;11(5):709–22.

85. Patel TK, Thakkar SH, Sharma D. Cutaneous adverse drug reactions in Indian population: A systematic review. Indian Dermatol Online J. 2014;5(Suppl 2):S76–86.

86. Porebski G, Pecaric-Petkovic T, Groux-Keller M, Bosak M, Kawabata T, Pichler W. In vitro drug causality assessment in Stevens-Johnson syndrome - alternatives for lymphocyte transformation test. Clin Exp Dermatol. 2013;43(9):1027–37.

87. Puavilai S, Noppakun N, Sitakalin C, Leenutaphong V, Wattanakrai P, Nakakes A, et al. Drug eruptions at five institutes in Bangkok. J Med Assoc Thail. 2005;88(11):1642–50.

88. Renda F, Landoni G, Bertini Malgarini R, Assisi A, Azzolini ML, Mucchetti M, et al. Drug reaction with eosinophilia and systemic symptoms (DRESS): A national analysis of data from 10-year post-marketing surveillance. Drug Saf. 2015;38(12):1211–8.

89. Rodríguez-Martín S, Martín-Merino E, Lerma V, Rodríguez-Miguel A, González O, González-Herrada C, et al. Active surveillance of severe cutaneous adverse reactions: A case-population approach using a registry and a health care database. Pharmacoepidemiol Drug Saf. 2018;27(9):1042–50.

90. S RB, Narayan SS, Sharma G, Rodrigues RJ, Kulkarni C. Pattern of adverse drug reactions to anti-epileptic drugs: A cross-sectional one-year survey at a tertiary care hospital. Pharmacoepidemiol Drug Saf. 2008;17(8):807–12.

91. Roy D, Purkayastha A, Tigga R. Analysis of adverse drug reaction in a tertiary care hospital: A retrospective study. Asian J Pharm Clin Res. 2017;10(1):347–9.

92. Saha A, Das N, Hazra A, Gharami R, Chowdhury S, Datta P. Cutaneous adverse drug reaction profile in a tertiary care out patient setting in Eastern India. Indian J Pharmacol. 2012;44(6):792– 7.

93. Sasidharanpillai S, Riyaz N, Khader A, Rajan U, Binitha MP, Sureshan DN. Severe cutaneous adverse drug reactions: A clinicoepidemiological study. Indian J Dermatol. 2015;60(1):102.

94. Sassolas B, Haddad C, Mockenhaupt M, Dunant A, Liss Y, Bork K, et al. ALDEN, an algorithm for assessment of drug causality in Stevens-Johnson syndrome and toxic epidermal necrolysis: Comparison with case-control analysis. Clin Pharmacol Ther. 2010;88(1):60–8.

95. Shalayel MHF, Ayed IAM, Huneif MA, Kordofani YM. A retrospective evaluation of cutaneous adverse drug reactions (CADRs) due to antibiotics using Naranjo adverse drug reactions (ADRs) probability scale. J Young Pharm. 2018;10(1):113–6.

96. Sharma V, Sethuraman G, Kumar B. Cutaneous adverse drug reactions: clinical pattern and causative agents--a 6 year series from Chandigarh, India. J Postgrad Med. 2001;47(2):95–9.

97. Sharma VK, Sethuraman G, Minz A. Stevens Johnson syndrome (SJS), toxic epidermal necrolysis (TEN) and SJS-TEN overlap: A retrospective study of causative drugs and clinical outcome. Indian J Dermatol Venereol Leprol. 2008;74(3):238–40.

98. Sharma R, Dogra D, Dogra N. A study of cutaneous adverse drug reactions at a tertiary center in Jammu, India. Indian Dermatol Online J. 2015;6(3):168–71.

99. Sheth H, Patel R, Chaudhary R, Malhotra S. Analysis of cutaneous adverse drug reactions presenting to the dermatology department of a tertiary care teaching hospital: A prospective, observational study. Int J Pharm Sci Res. 2019;10(3):1253–8.

100. Sim DW, Yu JE, Jeong J, Jung J-W, Kang H-R, Kang DY, et al. Variation of clinical manifestations according to culprit drugs in DRESS syndrome. Pharmacoepidemiol Drug Saf. 2019;28(6):840–8.

101. Son Y-M, Lee J-R, Roh J-Y. Causality assessment of cutaneous adverse drug reactions. Ann Dermatol. 2011;23(4):432–8.

102. Sousa-Pinto B, Araújo L, Freitas A, Correia O, Delgado L. Stevens-Johnson syndrome/toxic epidermal necrolysis and erythema multiforme drug-related hospitalisations in a national administrative database. Clin Transl Allergy. 2018;8(2).

103. Su P, Aw CWD. Severe cutaneous adverse reactions in a local hospital setting: A 5-year retrospective study. Int J Dermatol. 2014;53(11):1339–45.

104. Sudershan V, Siddiqua S, Aruna D, Manmohan, Ramesh S, Yasmeen N. Cutaneous adverse drug reactions in a tertiary care hospital. Der Pharm Lett. 2012;4(2):408–13.

105. Sushma M, Noel M, Ritika M, James J, Guido S. Cutaneous adverse drug reactions: a 9-year study from a South Indian Hospital. Pharmacoepidemiol Drug Saf. 2005;14(8):567–70.

106. Talebi R, Saki N, Raeisi Shahraki H, Owji SH. An epidemiological study of Stevens-Johnson syndrome and toxic epidermal necrolysis during 2010-2015 at Shahid Faghihi Hospital, Shiraz, Iran. Iran J Med Sci. 2018;43(4):421–5.

107. Tat L, Chiat T, Ying Y, Muniandy P. Steven-Johnson syndrome and toxic epidermal necrolysis in Northern Sarawak between year 2011 and 2015: A retrospective review of causative agents and clinical outcome. J Pakistan Assoc Dermatologists. 2017;27(3):243–6.

108. Thakkar S, Patel TK, Vahora R, Bhabhor P, Patel R. Cutaneous Adverse Drug Reactions in a Tertiary Care Teaching Hospital in India: An Intensive Monitoring Study. Indian J Dermatol. 2017;62(6):618–25.

109. Tripathy R, Pattnaik KP, Dehury S, Patro S, Mohanty P, Sahoo SS, et al. Cutaneous adverse drug reactions with fixed-dose combinations: Special reference to self-medication and preventability. Indian J Pharmacol. 2018;50(4):192–6.

110. Wang F, Zhao Y-K, Li M, Zhu Z, Zhang X. Trends in culprit drugs and clinical entities in cutaneous adverse drug reactions: a retrospective study. Cutan Ocul Toxicol. 2017;36(4):370–6.

111. Wang L, Mei X-L. Retrospective Analysis of Stevens-Johnson Syndrome and Toxic Epidermal Necrolysis in 88 Chinese Patients. Chin Med J (Engl). 2017;130(9):1062–8.

112. Wang L, Mei X-L. Drug Reaction with Eosinophilia and Systemic Symptoms: Retrospective Analysis of 104 Cases over One Decade. Chin Med J (Engl). 2017;130(8):943–9.

113. Wang Y-H, Chen C-B, Tassaneeyakul W, Saito Y, Aihara M, Choon SE, et al. The Medication Risk of Stevens-Johnson Syndrome and Toxic Epidermal Necrolysis in Asians: The Major Drug Causality and Comparison With the US FDA Label. Clin Pharmacol Ther. 2019;105(1):112–20.

114. Waton J, Trechot P, Loss-Ayav C, Schmutz JL, Barbaud A. Negative predictive value of drug skin tests in investigating cutaneous adverse drug reactions. Br J Dermatol. 2009;160(4):786–94.

115. Wolfson A, Zhou L, Li Y, Blumenthal K. Drug rash eosinophilia and systemic symptoms (dress) syndrome identified in electronic health record allergy module. Ann Allergy, Asthma Immunol. 2017;119(5):S1.

116. Yang M-S, Lee J, Kim J, Kim G-W, Kim B-K J.Y. K, et al. Searching for the Culprit Drugs for Stevens-Johnson Syndrome and Toxic Epidermal Necrolysis from a Nationwide Claim Database in Korea. J Allergy Clin Immunol Pract. 2020;8(2):690.

117. Yang C-Y, Dao R-L, Lee T-J, Lu C-W, Yang C-H, Hung S-I, et al. Severe cutaneous adverse reactions to antiepileptic drugs in Asians. Neurology. 2011;77(23):2025–33.

118. Yeung CK, Ma SY, Hon C, Peiris M, Chan HHL. Aetiology in sixteen cases of toxic epidermal necrolysis and Stevens-Johnson syndrome admitted within eight months in a teaching hospital. Acta Derm Venereol. 2003;83(3):179–82.

119. Zhao J, Hu L, Zhang L, Zhou M, Gao L, Cheng L. Causative drugs for drug-induced cutaneous reactions in central China: a 608-case analysis. An Bras Dermatol. 2019;94(6):664–70.

120. Behera SK, Das S, Xavier AS, Velupula S, Sandhiya S. Comparison of different methods for causality assessment of adverse drug reactions. Int J Clin Pharm. 2018 Aug 1;40(4):903–10.

121. Belhekar MN, Taur SR, Munshi RP. A study of agreement between the Naranjo algorithm and WHO-UMC criteria for causality assessment of adverse drug reactions. Indian J Pharmacol. 2014;46(1):117–20.

122. Gallagher RM, Kirkham JJ, Mason JR, Bird KA, Williamson PR, Nunn AJ, et al. Development and inter-rater reliability of the Liverpool adverse drug reaction causality assessment tool. PLoS One. 2011;6(12).

123. Koh Y, Shu CL. A new algorithm to identify the causality of adverse drug reactions. Drug Saf. 2005;28(12):1159–61.

124. Macedo AF, Marques FB, Ribeiro CF. Can decisional algorithms replace global introspection in the individual causality assessment of spontaneously reported ADRs? Drug Saf. 2006;29:697–702.

125. Mittal N, Gupta MC. Comparison of agreement and rational uses of the WHO and Naranjo adverse event causality assessment tools. J Pharmacol Pharmacother. 2015;6(2):91–3.

126. Mouton JP, Mehta U, Rossiter DP, Maartens G, Cohen K. Interrater agreement of two adverse drug reaction causality assessment methods: A randomised comparison of the Liverpool Adverse Drug Reaction Causality Assessment Tool and the World Health Organization-Uppsala Monitoring Centre system. PLoS One. 2017;12(2).

127. Thaker SJ, Sinha RS, Gogtay NJ, Thatte UM. Evaluation of inter-rater agreement between three causality assessment methods used in pharmacovigilance. J Pharmacol Pharmacother. 2016;7(1):31–3.

128. Varallo FR, Planeta CS, Herdeiro MT, De Mastroianni PC. Imputation of adverse drug reactions: Causality assessment in hospitals. PLoS One. 2017;12(2).

129. Aberer W. Drug hypersensitivities: The need for standardization. Eur Ann Allergy Clin Immunol. 2005;37(6):219–22.

130. Ardern-Jones MR, Mockenhaupt M. Making a diagnosis in severe cutaneous drug hypersensitivity reactions. Curr Opin Allergy Clin Immunol. 2019;19(4):283–93.

131. Ariza A, Torres MJ, Moreno-Aguilar C, Fernandez-Santamaria R, Fernandez TD. Early Biomarkers for Severe Drug Hypersensitivity Reactions. Curr Pharm Des. 2019;25(36):3829–39.

132. Barbaud A, Goncalo M, Bruynzeel D, Bircher A, Dermatitis ES of C. Guidelines for performing skin tests with drugs in the investigation of cutaneous adverse drug reactions. Contact Dermatitis. 2001;45(6):321–8.

133. Bergmann MM, Caubet J-C. Role of in vivo and in vitro Tests in the Diagnosis of Severe Cutaneous Adverse Reactions (SCAR) to Drug. Curr Pharm Des. 2019;25(36):3872–80.

134. Burbach GJ, Zuberbier T. Diagnosis of drug-induced skin reactions: a future role for computer-aided systems?. Curr Opin Allergy Clin Immunol. 2011;11(5):451–6.

135. Cabanas R, Ramirez E, Sendagorta E, Alamar R, Barranco R, Blanca-Lopez N, et al. Spanish Guidelines for Diagnosis, Management, Treatment and Prevention of DRESS syndrome. J Investig Allergol Clin Immunol. 2020;30(4):229–53.

136. Cacoub P, Musette P, Descamps V, Meyer O, Speirs C, Finzi L, et al. The DRESS syndrome: a literature review. Am J Med. 2011;124(7):588–97.

137. Daunton A, Farquharson N, Coulson I. Drug eruptions. Med (United Kingdom). 2017;45(7):422– 8.

138. Dodiuk-Gad RP, Chung W-H, Valeyrie-Allanore L, Shear NH. Stevens-Johnson Syndrome and Toxic Epidermal Necrolysis: An Update. Am J Clin Dermatol. 2015;16(6):475–93.

139. Dubey A, Prabhu S, Shankar P, Subish P, Prabhu M, Mishra P. Dermatological adverse drug reactions due to systemic medications - A review of literature. J Pakistan Assoc Dermatologists. 2006;16(1):28–38.

140. Ingen-Housz-Oro S, Duong T-A, Bensaid B, Bellon N, de Prost N, Lu D, et al. Epidermal necrolysis French national diagnosis and care protocol (PNDS; protocole national de diagnostic et de soins). Orphanet J Rare Dis. 2018;13(1):56.

141. Kotrulja L, Milavec-Puretic V, Pasic A. Diagnostic methods in the determination of drug allergy. Acta Dermatovenerologica Croat. 1997;5(3 PG-111–116):111–6.

142. Mereniuk A, Jaque A, Jeschke MG, Shear NH. Toxic Epidermal Necrolysis Spectrum Management at Sunnybrook Health Sciences Centre: Our Multidisciplinary Approach After Review of the Current Evidence. J Cutan Med Surg. 2018;22(2):213–9.

143. Patel TK, Barvaliya MJ, Sharma D, Tripathi C. A systematic review of the drug-induced Stevens-Johnson syndrome and toxic epidermal necrolysis in Indian population. Indian J Dermatol Venereol Leprol. 2013;79(3):389–98.

144. Paulmann M, Mockenhaupt M. Severe drug-induced skin reactions: clinical features, diagnosis, etiology, and therapy. J Dtsch Dermatol Ges. 2015;13(7):625–45.

145. Schopf E, Mockenhaupt M. Drug-attributability in severe skin reactions: Toxic epidermal necrolysis (TEN) and Stevens-Johnson syndrome (SJS). Korean J Dermatology. 1999;37(8):975– 82.

146. Sharma VK, Sethuraman G. Adverse cutaneous reactions to drugs: An overview. J Postgrad Med. 1996;42(1):15–22.

147. Meyboom RHB, Hekster YA, Egberts ACG, Gribnau FWJ, Edwards IR. Causal or casual? The role of causality assessment in pharmacovigilance. Drug Saf. 1997;17(6):374–89.

148. Meyboom RHB. Causality assessment revisited. Pharmacoepidemiol Drug Saf. 1998;7(S1):S63-5.

149. Pande S. Causality or Relatedness assessment in adverse drug reaction and its relevance in dermatology. Indian J Dermatol. 2018;63(1):18–21.

150. Ralph Edwards I. Causality Assessment in Pharmacovigilance: Still a Challenge. Drug Saf. 2017;40(5):365–72.

